# Efficacy and safety of potassium-containing low-sodium salt substitutes for cardiovascular disease prevention in mixed adult populations: an umbrella review

**DOI:** 10.64898/2026.05.06.26352501

**Authors:** Kirstin P. West, Kokou Tbah Tighankpa, Zikang Fang, Mamadou Moustapha Ndiaye, Natalie Zhang, Serena Chu, Sophia Li, Amanda Shiach, Nicole S. Dettmar, Adam Drewnowski, Yanfang Su

## Abstract

Potassium-containing low-sodium salt substitutes (LSSS) may lower sodium intake, increase potassium intake, and reduce cardiovascular risk in mixed adult populations, but the review literature is overlapping and methodologically heterogeneous. This umbrella review assessed the efficacy, safety, and evidence quality of potassium-containing LSSS for blood pressure, cardiovascular outcomes, and adverse events. Following a registered PROSPERO protocol (CRD420261294404), we searched PubMed, Embase, Web of Science, Global Health (EBSCO) and the Cochrane Database of Systematic Reviews from inception to 6 March 2026 for systematic reviews, meta-analyses and umbrella reviews of potassium-containing LSSS. Eleven reviews met eligibility criteria. Methodological confidence was high in one review, moderate in three, low in five and critically low in two. Primary-study overlap was very high (corrected covered area 28.5%). Review-level pooled estimates consistently favoured potassium-containing LSSS for systolic blood pressure (mean differences −4.61 to −8.87 mmHg) and diastolic blood pressure (−1.42 to −4.04 mmHg). Later reviews also reported lower all-cause mortality (RR 0.88–0.89), cardiovascular mortality (RR 0.72–0.87), composite cardiovascular events and selected stroke outcomes; however, clinical-outcome estimates were heavily influenced by the Salt Substitute and Stroke Study. Serum potassium changed minimally (−0.02 to 0.18 mmol/l), and pooled estimates for hyperkalaemia and serious adverse events showed no clear excess risk in trial populations that largely excluded participants at higher risk of impaired potassium handling. Potassium-containing LSSS consistently lower blood pressure and may improve cardiovascular outcomes, but further trials are needed outside Eastern Asia, with clearer formulation reporting, prespecified baseline CVD-history strata, and stronger safety data in higher-risk populations.

## Introduction

Cardiovascular disease (CVD) remains the leading cause of death globally.^[1,2]^ The World Health Organization (WHO) estimates that 19.8 million deaths were attributable to CVD in 2022, representing approximately 32% of all global deaths.^[3]^ Elevated blood pressure is a major modifiable driver of myocardial infarction, stroke, heart failure, and cardiovascular mortality.^[4–7]^ From a nutrition and public health perspective, excess sodium intake and insufficient potassium intake remain among the most important dietary determinants of raised blood pressure and downstream cardiovascular risk.^[4,8]^

Population sodium intake remains far above recommended levels. WHO recommends that adults consume less than 2 g sodium per day (approximately 5 g salt/day) and at least 3.51 g potassium per day.^[9,10]^ However, the 2025 WHO guideline on lower-sodium salt substitutes (LSSS) cites an estimated mean global sodium intake of approximately 4.3 g/day (10.78 g/day salt), more than double the recommended threshold.^[11,12]^ This gap is especially important in settings where discretionary salt added during home cooking or at the table contributes substantially to total sodium intake. In such contexts, sustained dietary change through advice alone can be difficult to achieve, and poor adherence to sodium-reduction recommendations remains an important barrier to the prevention of hypertension and stroke.^[7,11,13]^

Potassium-containing LSSS replace part of sodium chloride (NaCl) with potassium chloride (KCl), thereby reducing sodium exposure while increasing potassium exposure.^[14,15]^ This is an attractive nutritional intervention because it can improve dietary electrolyte balance without requiring major changes in cooking practices or food choice.^[13,14]^ In January 2025, WHO issued a conditional recommendation supporting replacement of regular table salt with potassium-containing LSSS for adults in general populations, while excluding pregnant women, children, people with kidney impairment and others with conditions or medication use that may compromise potassium excretion.^[15]^ Potassium-containing LSSS therefore has clear policy relevance: they are simple, food-based and potentially scalable, but their efficacy, generalisability and safety profile require careful interpretation. Real-world implementation, however, is likely to depend not only on efficacy and safety, but also on consumer awareness, acceptability, product availability, and clear communication about groups in whom potassium-enriched products may be inappropriate.^[15–18]^

The evidence base has expanded rapidly, with randomised trials, pairwise meta-analyses and network meta-analyses reporting potassium-containing LSSS benefits for blood pressure and, in some settings, stroke, composite cardiovascular events and mortality.^[1,19–25]^ However, the review literature is now overlapping and heterogeneous. Existing syntheses vary in the populations included, the LSSS formulations compared, the outcomes emphasised and the extent to which safety outcomes are assessed.^[19–25]^ Several newer mortality and cardiovascular-event estimates are also heavily influenced by a small number of large trials, particularly the Salt Substitute and Stroke Study (SSaSS).^[19,24]^ Without a review-of-reviews approach, it is difficult to determine which syntheses should anchor interpretation and where uncertainty remains.^[26–29]^

An umbrella review is therefore needed to consolidate review-level evidence on the efficacy and safety of potassium-containing LSSS for cardiovascular prevention in mixed adult populations, evaluate methodological quality, and quantify the overlap across primary studies.^[26–29]^ Our review was guided by an a-priori registered protocol^[30]^ and organised around a causal framework linking LSSS exposure to validated intake biomarkers, blood-pressure responses, clinical cardiovascular outcomes as reported by source reviews, and safety outcomes (**Figure 1**).^[6,14,15]^ We planned to distinguish primary- and secondary-prevention populations, but most source reviews and underlying trials did not stratify by prior CVD history. We therefore treated the available evidence as mixed or unclear adult cardiovascular-prevention populations and extracted blood-pressure status, age and other subgroup information where reported. We also considered implementation-relevant outcomes, including acceptability, palatability and feasibility, because these influence whether a nutritionally effective intervention can achieve population impact in real-world settings.^[13,18,31,32]^

**Figure 1.**
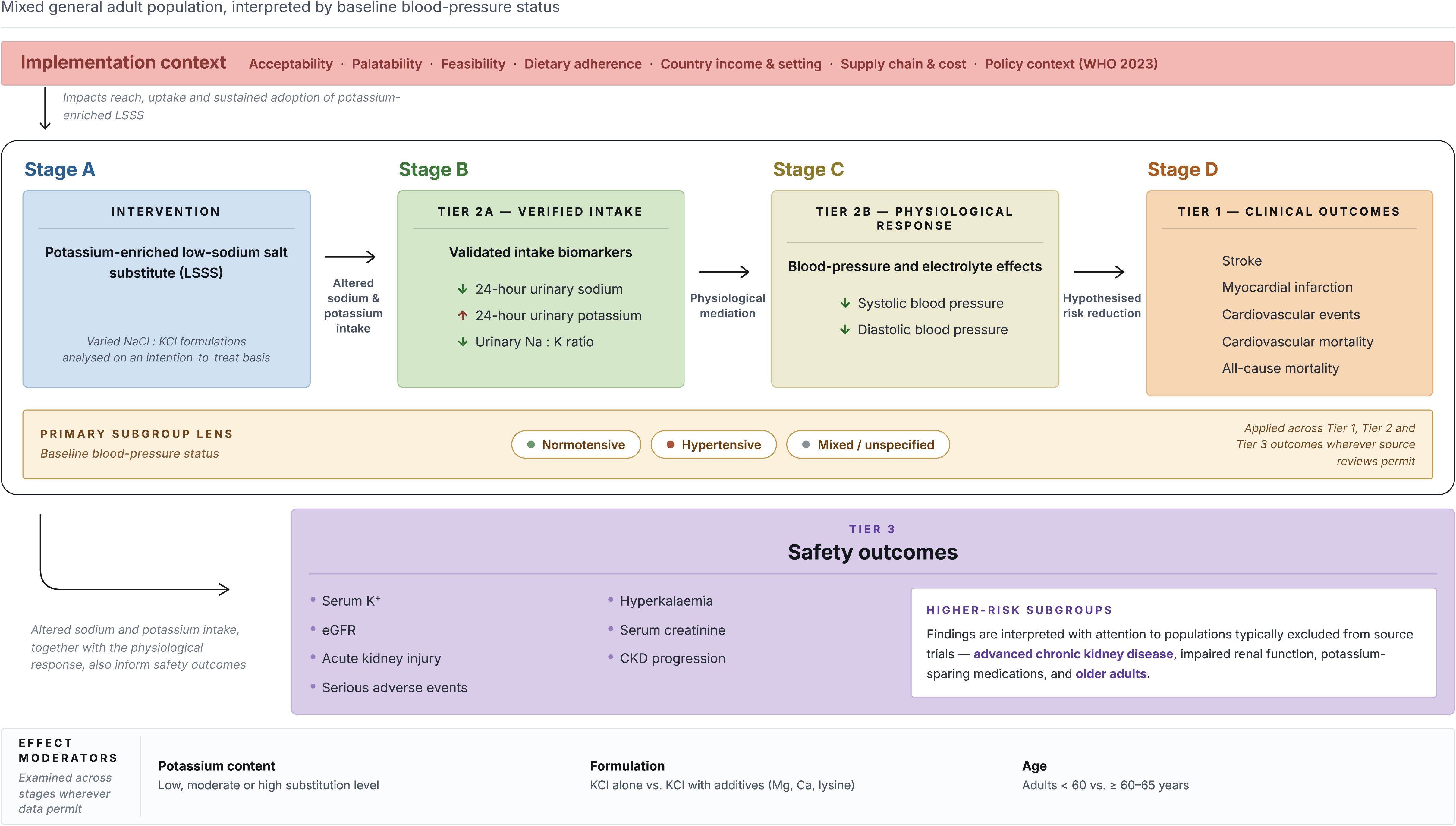
Conceptual causal pathway and safety framework for potassium-containing low-sodium salt substitutes (LSSS). Potassium-containing LSSS replace part of sodium chloride (NaCl) with potassium chloride (KCl), thereby reducing sodium exposure while increasing potassium exposure at the point of use. The hypothesised efficacy pathway proceeds from intervention/exposure (Stage A) to validated intake biomarkers, including 24-h urinary sodium, 24-h urinary potassium and urinary sodium-to-potassium (Na:K) ratio (Stage B; Tier 2a), then to physiological response in systolic and diastolic blood pressure (Stage C; Tier 2b), and finally to clinical endpoints, including stroke, major adverse cardiovascular events (MACE), cardiovascular mortality and all-cause mortality (Stage D; Tier 1). Safety outcomes (Tier 3), including serum potassium, hyperkalaemia and renal outcomes (e.g. eGFR, creatinine, AKI and CKD), are assessed in parallel because increases in potassium intake may be inappropriate in individuals with impaired potassium excretion or other conditions affecting potassium handling. Abbreviations: LSSS, low-sodium salt substitute; SBP, systolic blood pressure; DBP, diastolic blood pressure; Na:K, sodium-to-potassium ratio; eGFR, estimated glomerular filtration rate; AKI, acute kidney injury; CKD, chronic kidney disease; CVD, cardiovascular disease.

Accordingly, this umbrella review aimed to assess the efficacy, safety, and evidence quality of potassium-containing LSSS for CVD prevention in mixed adult populations, with attention to review quality, primary-study overlap, blood pressure status, formulation heterogeneity, and safety-related subgroups.

## Methods

### Protocol and registration

This umbrella review was prospectively registered in PROSPERO (<u>CRD420261294404</u>).^[30]^ Reporting was informed by PRISMA 2020^[33]^, the PRIOR statement for overviews of reviews, and umbrella-review methodological guidance.^[26–29]^

### Eligibility criteria

Eligible records were systematic reviews with or without meta-analysis, meta-analyses, and umbrella reviews that included at least one primary study evaluating a potassium-containing low-sodium salt substitute and reported a reproducible search and study-selection process. The intervention of interest was potassium-containing LSSS used to replace regular salt in cooking, seasoning, food preparation, or food reformulation to reduce dietary sodium intake and increase potassium intake. For eligibility, the substitute had to contain potassium chloride as a sodium-replacing constituent; products without potassium chloride were excluded. Because formulation reporting was heterogeneous across reviews, we did not prespecify a minimum potassium chloride percentage but extracted reported NaCl:KCl compositions where available. Eligible comparators were regular salt, usual care, no intervention, or a placebo (e.g., no salt substitute).

We excluded narrative or non-systematic reviews, editorials, commentaries, protocols, conference abstracts without sufficient review data, and single primary studies because the umbrella review aimed to synthesise reproducibly identified review-level evidence. Detailed eligibility criteria and category-level justification of excluded record/study types are provided in **Supplementary Table S1**. We excluded reviews covering interventions with general sodium-reduction advice or dietary patterns without a salt-substitute component, as well as potassium supplementation pills and non-potassium salt alternatives such as monosodium glutamate and herb blends. Broad sodium-reduction policy reviews were also excluded unless extractable data specific to potassium-containing LSSS were reported. No restrictions were applied regarding language.

### Information sources and search strategy

The librarian author (N.S.D) conducted an electronic literature search up to 6 March, 2026, for published studies using five major databases: PubMed, Embase, Web of Science, Global Health (EBSCO) and Cochrane (Systematic Reviews only). These databases were predefined in the preregistered PROSPERO protocol (<u>CRD420261294404</u>).^[30]^ A total of 839 records were imported into Covidence, and duplicates were removed both automatically by Covidence and manually by the reviewers.^[34]^ Detailed search strategies for each database are presented in the **Supplementary Table S2**. Records published after the search cut-off date were not eligible for inclusion, in accordance with the preregistered protocol.

### Study selection

Study selection was performed in Covidence in two stages. In the first phase, two independent reviewers (K.P.W. and K.T.T.) screened titles and abstracts. In the second phase, the full texts of potentially relevant articles were uploaded to Covidence and independently evaluated by the same reviewers.^[34]^ Before full text screening, the two reviewers completed a pilot calibration exercise on the first 50 records to harmonise interpretation of the eligibility criteria and refine the screening approach. Disagreements at any stage were resolved through discussion, and when necessary, adjudicated by a third reviewer (Y.S.).

### Data extraction

Data from the 11 included reviews were extracted independently and in duplicate in Covidence by reviewer pairs drawn from K.P.W., N.Z., S.C., S.L. and Z.F. using a standardised extraction form.^[34]^ Extracted items included bibliographic characteristics, review type, search sources and last search date, included study designs, number of included studies and participants, settings, geographic scope, population descriptors including blood-pressure status and CVD history where reported, potassium-containing LSSS formulation, comparator(s), outcomes assessed, and reported effect measures and summary estimates. We also recorded reporting characteristics relevant to appraisal of the review literature, including protocol registration, PRISMA adherence^[33]^, performance of meta-analysis, certainty-of-evidence assessment using GRADE^[35,36]^, and assessment of publication bias. Discrepancies between extractors were resolved by discussion and, when required, by senior-reviewer adjudication (Y.S.). After reconciliation of the two human extractions, Claude.ai (Claude Sonnet 4.6, Anthropic, PBC, https://claude.ai, accessed March 2026) was used as a secondary consistency-checking tool to compare the full-text report against the prespecified extraction schema and flag potentially missing or discordant items. All AI-flagged items were checked against the source report by a human reviewer before any change was accepted. AI outputs were not used to determine eligibility, final extracted values, AMSTAR 2 or GRADE judgements, effect estimates or conclusions.

### Assessment of methodological quality, overlap and certainty of evidence

Methodological quality of included reviews was appraised independently by two reviewers drawn from K.P.W., N.Z., S.C., S.L. and Z.F. using AMSTAR 2, with item-level judgements and overall confidence ratings recorded for each review.^[37]^ Disagreements were resolved by consensus, with senior-reviewer adjudication when needed (Y.S.). To address redundancy across the review literature, we mapped primary studies in a citation matrix and quantified overlap using corrected covered areas (CCA).^[26]^ When multiple reviews addressed the same question with substantial overlap, we did not combine pooled estimates across reviews. Instead, interpretive emphasis was placed on the most recent and methodologically robust review that most directly addressed the outcome and population descriptor of interest, while the range of effect estimates across overlapping reviews was also reported. If de novo synthesis of primary randomised trials had been required to clarify conflicting or incomplete review-level evidence, risk of bias would have been assessed independently by two reviewers using the Cochrane Risk of Bias 2 tool, including the cluster-randomised version where applicable.^[38,39]^ Certainty of evidence for any such de novo synthesis was assessed with GRADE across the domains of risk of bias, inconsistency, indirectness, imprecision, and publication bias.^[35,36]^

### Summary measures

Effect measures were extracted as reported by the included reviews. For dichotomous outcomes, these included risk ratios, rate ratios, odds ratios, hazard ratios, or absolute effects when reported. For continuous outcomes, these included mean differences or standardised mean differences with corresponding 95% confidence intervals and reported units. We also extracted the number of contributing studies and participants, follow-up duration, heterogeneity statistics (e.g. I²)^[40]^, and any certainty assessments reported by review authors. Outcomes were grouped a priori according to the prespecified umbrella-review framework as Tier 1 clinical outcomes (e.g. stroke, myocardial infarction or acute coronary syndrome where available, cardiovascular mortality, all-cause mortality and composite cardiovascular events as defined by source reviews), Tier 2 validated intermediate outcomes (e.g. systolic and diastolic blood pressure, 24-h urinary sodium excretion, 24-h urinary potassium excretion, urinary sodium-to-potassium ratio), Tier 3 safety outcomes (e.g. serum potassium, hyperkalaemia, renal outcomes, serious adverse events), and exploratory implementation outcomes (e.g. acceptability, palatability and feasibility).

### Data synthesis and analysis

Evidence was synthesised primarily at the review level and grouped by outcome tier, population descriptors, review design, and reported blood-pressure status where available. Reviews that did not report results separately by prior CVD history were classified as mixed or unclear rather than as primary or secondary prevention. Because meta-analysing pooled estimates across overlapping reviews would risk double-counting the same primary studies, review-level pooled estimates were not combined when substantial overlap was present. Instead, we summarised the direction, magnitude, precision, heterogeneity and certainty of findings across reviews, with greater weight given to a prespecified anchor review selected based on recency, AMSTAR 2 confidence, completeness of reporting, and direct relevance to the outcome under consideration. De novo pooling of primary randomised trials was considered only when review-level evidence was materially discordant, CVD-history-specific estimates could not be recovered from the included reviews, or a clinically important outcome remained incompletely synthesised despite extractable primary-trial data. Safety and implementation outcomes were summarised narratively. Where undertaken, de novo meta-analysis used random-effects models and prespecified subgroup analyses when feasible.^[41]^ Citation mapping overlap calculations, quantitative summaries and base figure outputs were undertaken in R (version 4.5.3; R Core Team, 2026).^[42]^ Visual layout refinements for R-generated figures were made using Claude Design (Claude Sonnet 4.6, Anthropic, PBC, https://claude.ai, accessed April 2026).

## Results

### Study selection

Database searches identified 839 records, including 10 records from the March 2026 update. After removal of 208 duplicates, 631 records underwent title and abstract screening, of which 548 were excluded. Eighty-three full-text articles were assessed for eligibility. Seventy-two articles were excluded, most commonly because the intervention was not a potassium-containing low-sodium salt substitute or LSSS-specific data were not extractable (n=35), the review type was ineligible (n=14), or the publication was abstract-only (n=12). Eleven reviews met the eligibility criteria and were included in the umbrella review (**Figure 2**).^[20–25,43–47]^

**Figure 2.**
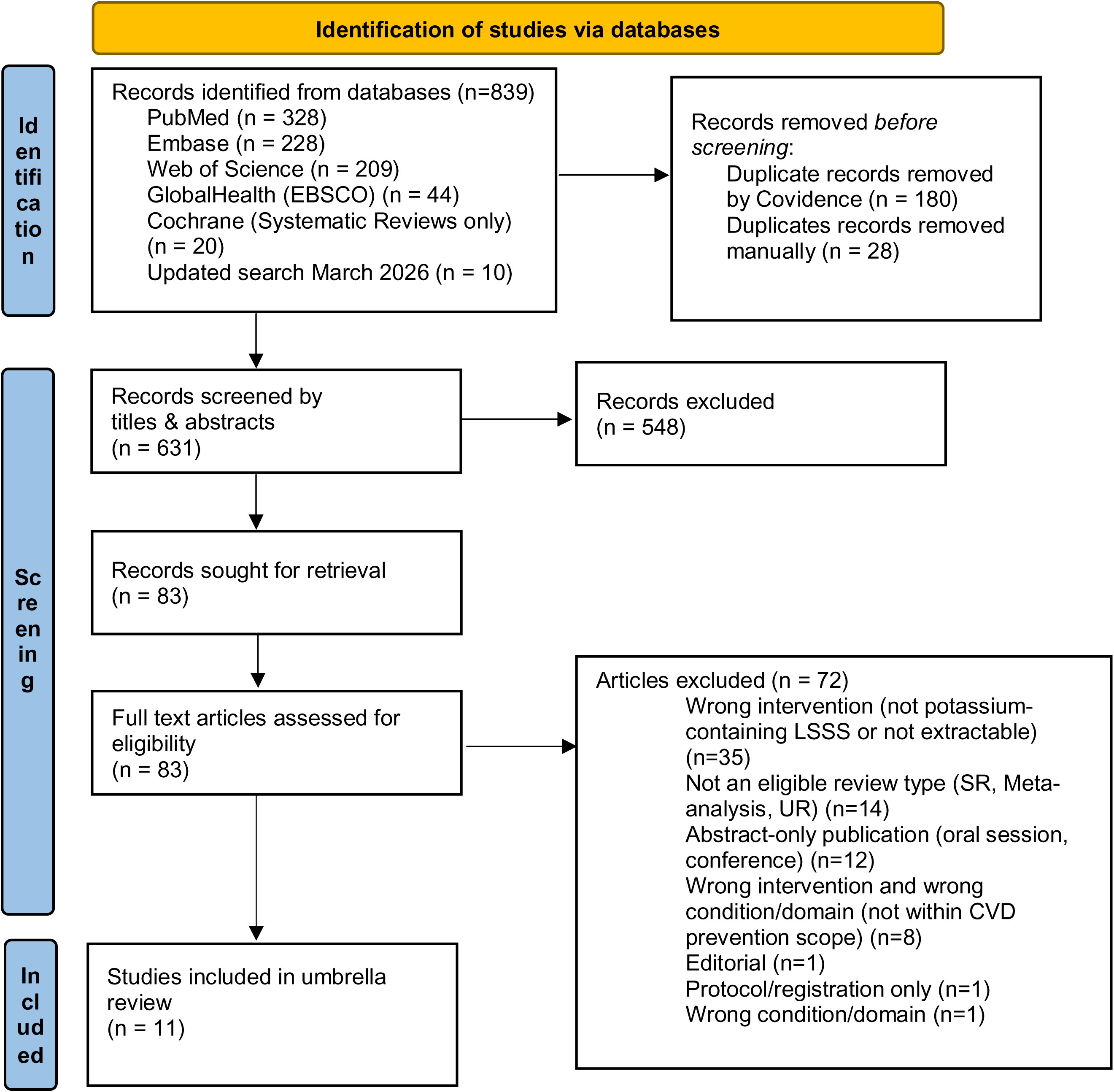
PRISMA 2020 flow diagram of record identification, screening, eligibility assessment and inclusion of reviews in the umbrella review. Flow diagram showing records identified across databases, duplicate removal, title and abstract screening, full-text eligibility assessment, reasons for exclusion and final inclusion of systematic reviews, meta-analyses and umbrella reviews. PRISMA Diagram Source: Page MJ, et al. BMJ 2021;372:n71. doi: 10.1136/bmj.n71. This work is licensed under CC BY 4.0. To view a copy of this license, visit https://creativecommons.org/licenses/by/4.0/

### Characteristics of included reviews

The eleven included reviews were published between 2019 and 2026 and comprised eight pairwise systematic reviews/meta-analyses and three network meta-analyses. Most synthesised randomised trials in adult populations and drew heavily on evidence from China and other Eastern Asia settings, usually in household, community, or institutional salt-replacement contexts. Seven reviews were specific to potassium-containing LSSS, whereas four broader syntheses contributed an extractable LSSS subgroup or network node. Outcome coverage broadened over time: earlier reviews focused mainly on blood pressure, whereas later reviews increasingly incorporated all-cause mortality, cardiovascular mortality, composite cardiovascular events, urinary biomarkers, and safety outcomes. Across reviews, formulation reporting, comparator choice, baseline CVD-history reporting, and outcome definitions were heterogeneous (**Supplementary Table S3**). Estimates for stroke, composite cardiovascular events, and mortality were repeatedly influenced by the Salt Substitute and Stroke Study^[19]^, particularly in later reviews reporting clinical outcomes.^[23–25,43]^

Outcome coverage was uneven across the prespecified three-tier framework. Tier 2 intermediate outcomes were the most consistently represented, with pooled systolic blood pressure (SBP) estimates available in ten reviews and pooled diastolic blood pressure (DBP) estimates in nine. All-cause mortality and cardiovascular mortality were each available as pooled relative estimates in four reviews, composite cardiovascular events in three reviews, and stroke subtype outcomes in one review. Tier 3 safety outcomes were concentrated mainly in serum potassium, whereas hyperkalaemia events, serious adverse events, serum creatinine, and adverse-event-related withdrawals were each available from single reviews only (**Supplementary Table S4**).

### Methodological quality of included reviews

Methodological quality varied across the 11 included reviews (**Figure 3**). One review was rated high confidence, three were rated moderate confidence, five low confidence and two critically low confidence. At the item level, duplicate study selection and duplicate data extraction were generally well addressed, and most reviews adequately described included studies and used appropriate statistical methods for meta-analysis. The most common limitations were absent or incomplete protocol registration due COVID-19 restrictions, lack of a study-level excluded-studies list with reasons, and incomplete reporting of funding sources for included primary studies. Moderate confidence under AMSTAR 2 indicates that a review has more than one non-critical weakness but no critical flaw; it should not be interpreted as meaning that the review is unreliable or low quality.^[37]^

**Figure 3.**
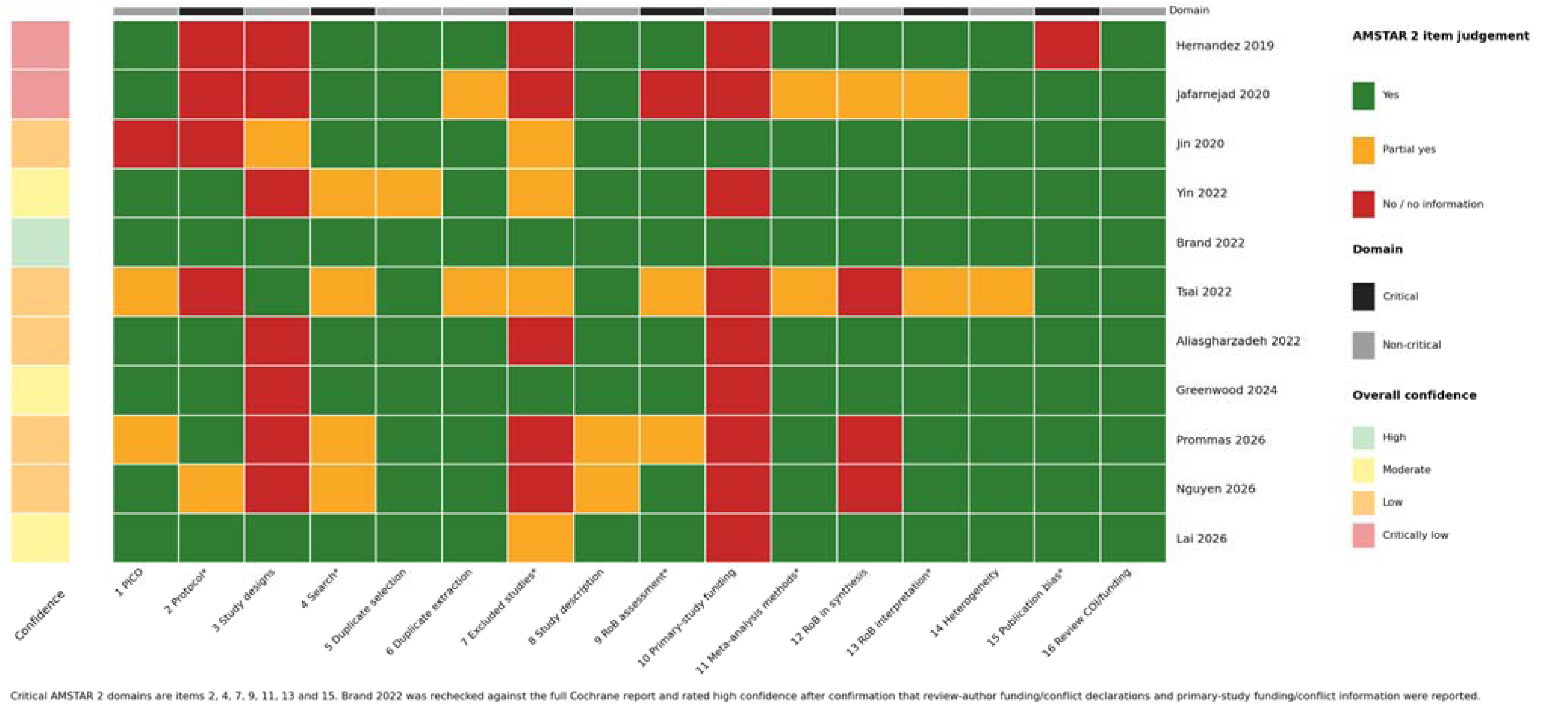
AMSTAR 2 item-level judgements across included reviews. AMSTAR 2 item-level judgement heatmap across included reviews. Heatmap showing ratings across all 16 AMSTAR 2 items for the 11 included reviews, with critical domains indicated separately from non-critical domains. Green = yes; yellow = partial yes; red = no or no information. Overall confidence ratings follow canonical AMSTAR 2 guidance and reflect the number and severity of critical and non-critical weaknesses.

### Primary study overlap across included reviews

Primary study overlap across included reviews was substantial (**Supplementary Figure S1; Supplementary Table S5**). The citation matrix comprised 154 review–study occurrences across 40 unique primary studies and 11 reviews, yielding an overall CCA of 28.5%, consistent with very high overlap. Thirty of the 40 unique primary studies (75%) appeared in more than one review, whereas only ten were represented in a single review. Across the 55 possible review pairs, 41 showed very high overlap (>15%), three high overlap (10 to <15%), four moderate overlap (5 to <10%) and seven slight overlap (<5%). The greatest pairwise overlap was observed for Brand et al. 2022 versus Lai et al. 2026 (CCA 57.1%), Yin et al. 2022 versus Tsai et al. 2022^[21]^ (56.5%), and Hernandez et al. 2019 versus Yin et al. 2022 (56.5%). Lower-overlap nodes were concentrated in pairs involving Nguyen et al. 2026.

### Review-level outcomes

#### Tier 1 clinical outcomes

Five review-level estimates were available for all-cause mortality. Point estimates were tightly clustered, ranging from RR 0.88 to 0.89, but precision differed across reviews. Hernandez et al. 2019 reported a non-significant estimate (RR 0.89, 95% CI 0.77 to 1.03; k=2; n=2159), whereas Yin et al. 2022, Brand et al. 2022, Tsai et al. 2022, and Greenwood et al. 2024 each reported estimates below 1.00 with confidence intervals excluding the null. Reported estimates were RR 0.89 (95% CI 0.85 to 0.94) for Yin et al. 2022, RR 0.89 (95% CI 0.83 to 0.95) for Brand et al. 2022, HR 0.88 (95% CI 0.82 to 0.94) for Tsai et al. 2022, and RR 0.88 (95% CI 0.82 to 0.93) for Greenwood et al. 2024 (**Figure 4**).

**Figure 4.**
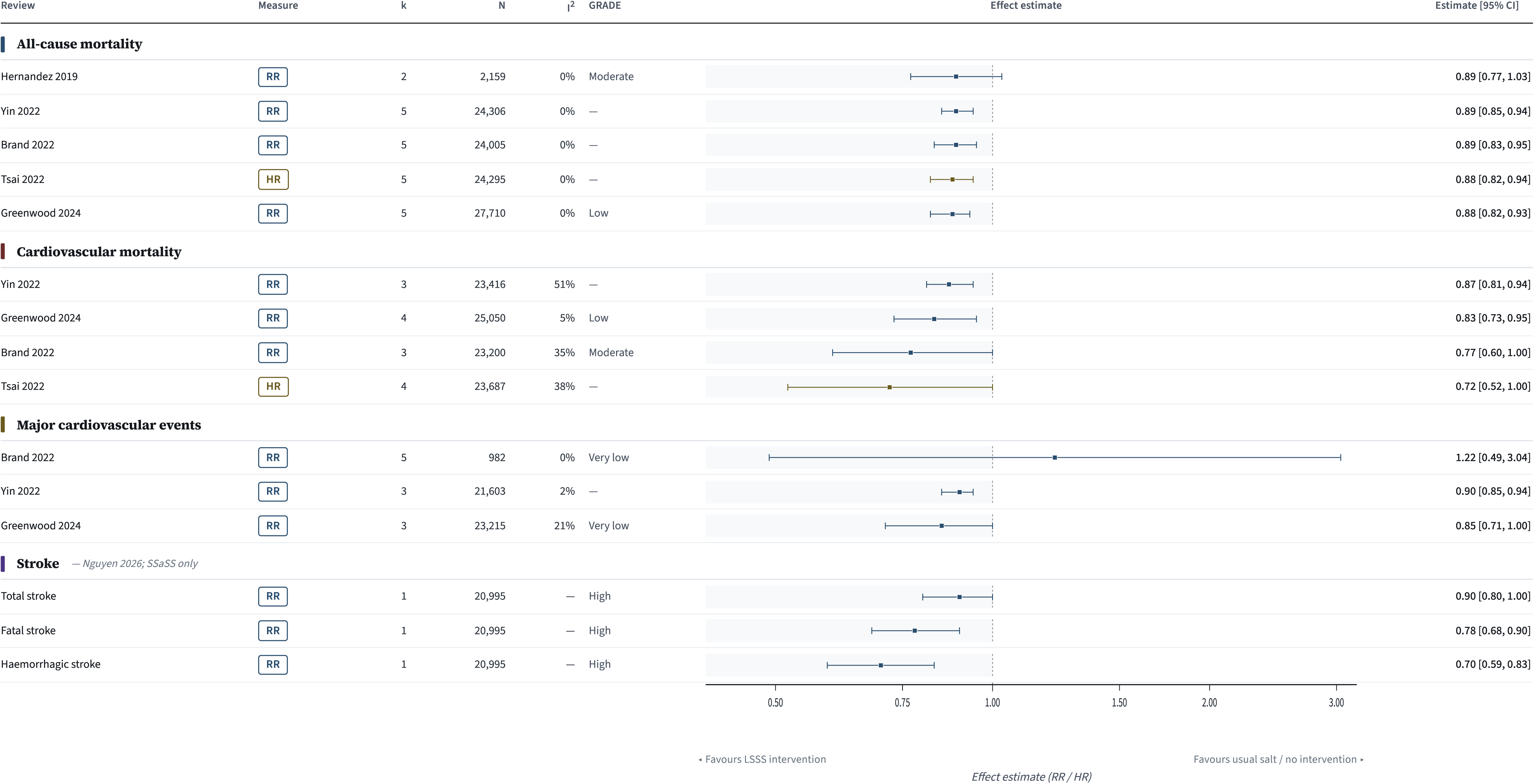
Review-level forest plots of Tier 1 clinical outcomes associated with potassium-containing low-sodium salt substitutes. Panels show pooled review-level estimates for all-cause mortality, cardiovascular mortality, major cardiovascular events and stroke outcomes, presented as reported by the source reviews and not pooled across reviews because of substantial primary-study overlap. Effect estimates below 1.00 favour potassium-containing low-sodium salt substitutes. Stroke outcomes from Nguyen 2026 are derived from a single direct trial (SSaSS). Where source reviews reported different effect measures (hazard ratio or rate ratio), estimates are displayed on the original reported scale.

Four review-level estimates were available for cardiovascular mortality. These ranged from 0.72 to 0.87. Yin et al. 2022 reported RR 0.87 (95% CI 0.81 to 0.94), Greenwood et al. 2024 reported RR 0.83 (95% CI 0.73 to 0.95), Brand et al. 2022 reported a RR 0.77 (95% CI 0.60 to 1.00), and Tsai et al. 2022 reported HR 0.72 (95% CI 0.52 to 1.00). Thus, all point estimates were below 1.00, although the Brand 2022 and Tsai 2022 confidence intervals extended to include 1.00 (**Figure 4**).

Three review-level estimates were available for major or composite cardiovascular events. Yin et al. 2022 reported RR 0.90 (95% CI 0.85 to 0.94) and Greenwood et al. 2024 reported RR 0.85 (95% CI 0.71 to 1.00). Brand et al. 2022 reported RR 1.22 (95% CI 0.49 to 3.04), but this estimate was based on a heterogeneous cardiovascular composite rather than a standard major adverse cardiovascular event definition (**Figure 4**).

Stroke-specific pooled estimates were limited to Nguyen et al. 2026, in which the salt-substitute node was informed by a single trial, SSaSS. In that review, the estimate for total stroke was RR 0.90 (95% CI 0.80 to 1.00; k=1; n=20,995), the estimate for fatal stroke was RR 0.78 (95% CI 0.68 to 0.90), and the estimate for haemorrhagic stroke was RR 0.70 (95% CI 0.59 to 0.83) (**Figure 4**).

#### Tier 2 intermediate outcomes

Pooled intermediate-outcome estimates were available most consistently for blood pressure and urinary electrolytes (**Figure 5**). Ten reviews reported pooled SBP estimates for potassium-containing LSSS versus regular salt or usual salt, and all point estimates favoured potassium-containing LSSS. Mean differences ranged from −4.61 mmHg (Yin et al. 2022; 95% CI −6.07 to −3.14) to −8.87 mmHg (Jafarnejad et al. 2020; 95 % CI −11.19 to −6.55). The published salt-substitute network estimate from Prommas et al. 2026 was −6.78 mmHg (95 % CI −8.42 to −5.14). Nine reviews reported pooled DBP estimates, which also all favoured potassium-containing LSSS, with mean differences ranging from −1.42 mmHg (Lai et al. 2026; 95 % CI −2.57 to −0.26) to −4.04 mmHg (Jafarnejad et al. 2020; 95 % CI −5.70 to −2.39).

**Figure 5.**
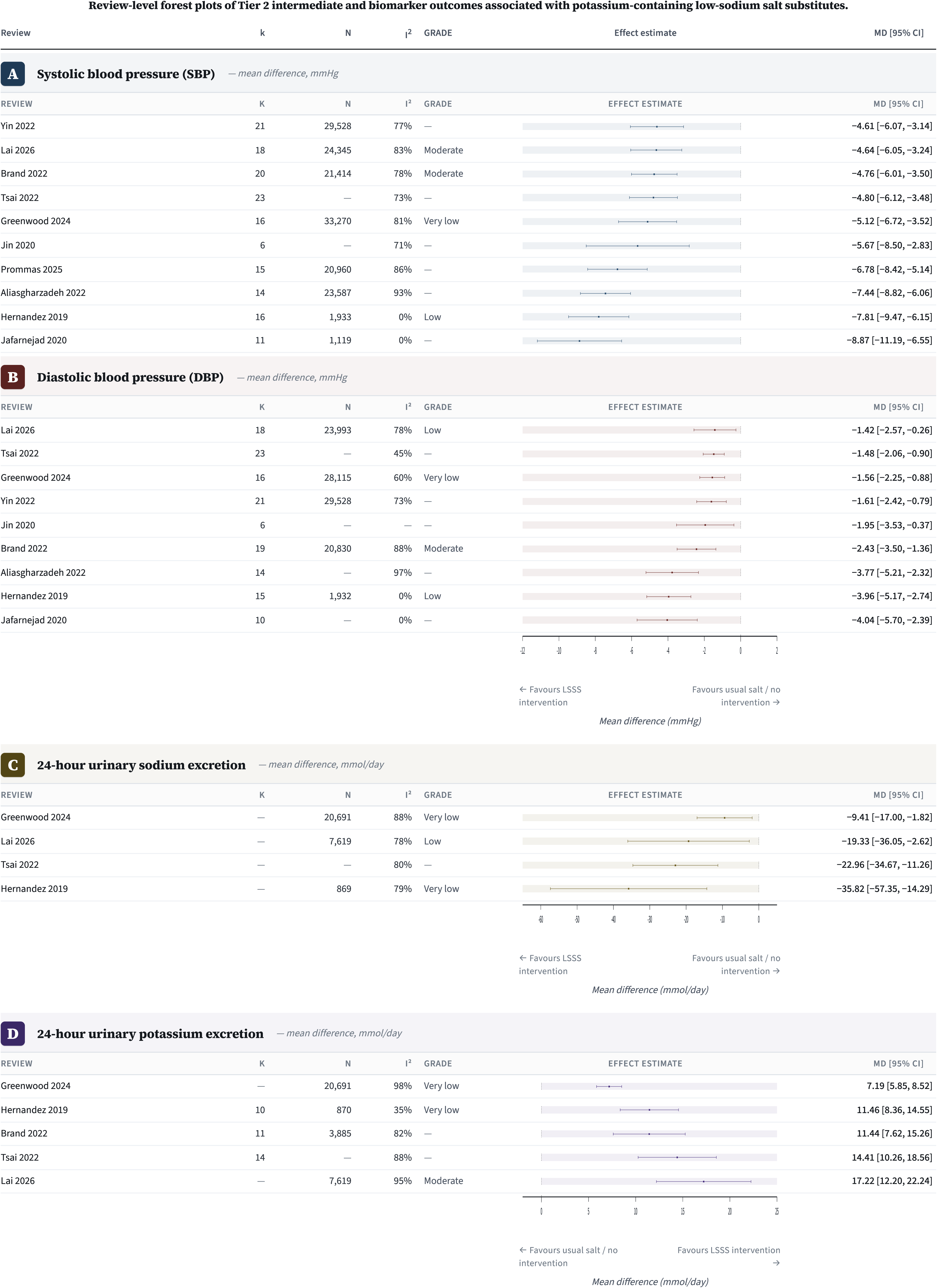
Review-level forest plots of Tier 2 intermediate and biomarker outcomes associated with potassium-containing low-sodium salt substitutes. Panels show pooled review-level estimates for systolic blood pressure, diastolic blood pressure, 24-h urinary sodium excretion and 24-h urinary potassium excretion. Negative mean differences for blood pressure and urinary sodium favour potassium-containing low-sodium salt substitutes; positive mean differences for urinary potassium favour potassium-containing low-sodium salt substitutes. Estimates are shown as reported by the source reviews and were not pooled across reviews because of substantial primary-study overlap. The urinary sodium-to-potassium ratio was pooled in one review only and is therefore reported in the text rather than as a separate panel.

Directly comparable pooled urinary sodium estimates were available from four reviews (**Figure 5**). All four estimates favoured potassium-containing LSSS, with mean differences of −35.82 mmol/d (Hernandez et al. 2019; 95 % CI −57.35 to −14.29), −22.96 mmol/d (Tsai et al. 2022; 95 % CI −34.67 to −11.26), −9.41 mmol/d (Greenwood et al. 2024; 95 % CI −17.00 to −1.82), and −19.33 mmol/d (Lai et al. 2026; 95 % CI −36.05 to −2.62). Directly comparable pooled urinary potassium estimates were available from five reviews, and all favoured potassium-containing LSSS, with mean differences of 11.46 mmol/d (Hernandez et al. 2019; 95 % CI 8.36 to 14.55), 11.44 mmol/d (Brand et al. 2022; 95 % CI 7.62 to 15.26), 14.41 mmol/d (Tsai et al. 2022; 95 % CI 10.26 to 18.56), 7.19 mmol/d (Greenwood et al. 2024; 95 % CI 5.85 to 8.52), and 17.22 mmol/d (Lai et al. 2026; 95 % CI 12.20 to 22.24). Brand et al. 2022 did not pool 24-h urinary sodium because heterogeneity was high, and Yin et al. 2022 reported urinary sodium and urinary potassium in g/d rather than mmol/d and was therefore not included in the directly comparable forest plots. Tsai et al. 2022 was the only included review to pool the 24-h urinary sodium-to-potassium ratio, reporting MD −0.63 (95 % CI −1.00 to −0.26).

#### Tier 3 safety outcomes

Five reviews reported pooled estimates for serum potassium (**Figure 6**). Point estimates ranged from −0.02 mmol/L (Yin et al. 2022; 95 % CI −0.49 to 0.44) to 0.18 mmol/L (Greenwood et al. 2024; 95 % CI 0.07 to 0.30). The remaining estimates were 0.12 mmol/L (Brand et al. 2022; 95 % CI 0.07 to 0.18), 0.10 mmol/L (Tsai et al. 2022; 95 % CI −0.02 to 0.23), and 0.11 mmol/L (Lai et al. 2026; 95 % CI 0.07 to 0.15). Four out of the five point estimates were above zero. Pooled clinical safety outcomes were sparse. Brand et al. 2022 reported hyperkalaemia events only, with RR 1.04 (95 % CI 0.46 to 2.38; k=5). Greenwood et al. 2024 reported serious adverse events only, with RR 1.04 (95 % CI 0.87, 1.25).

**Figure 6.**
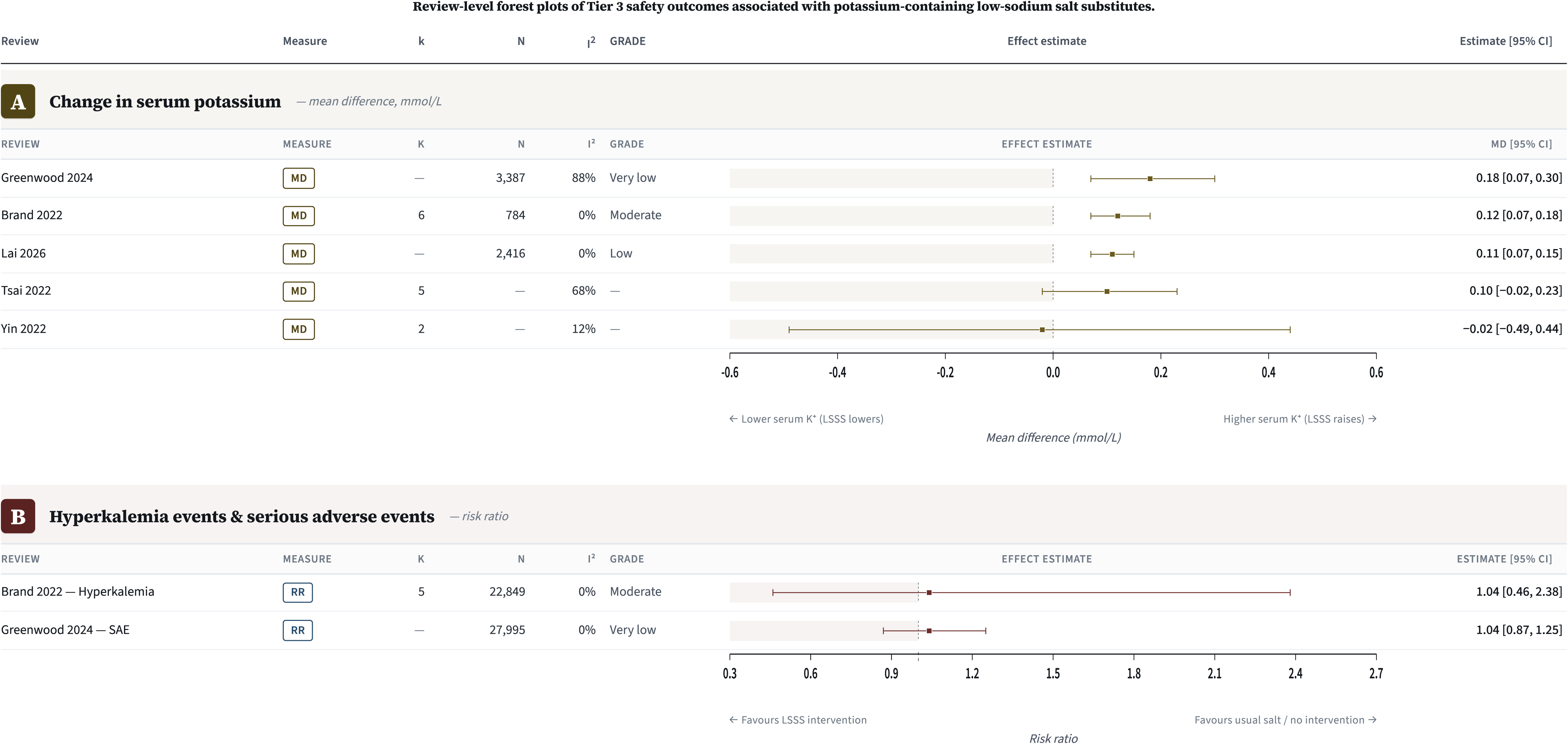
Review-level forest plots of Tier 3 safety outcomes associated with potassium-containing low-sodium salt substitutes. Panels show pooled review-level estimates for change in serum potassium and for hyperkalaemia events and serious adverse events. Positive mean differences indicate higher serum potassium with potassium-containing low-sodium salt substitutes; relative-effect estimates above 1.00 indicate higher event rates with potassium-containing low-sodium salt substitutes. Estimates are shown as reported by the source reviews and were not pooled across reviews because of substantial primary-study overlap. Included trial populations generally excluded participants with impaired potassium excretion or use of potassium-sparing medications.

#### Subgroup analyses by age and hypertensive status

Age-stratified SBP estimates were available in four reviews (**Supplementary Figure S2**). In Yin et al. 2022, Tsai et al. 2022, and Greenwood et al. 2024, the magnitude of the pooled SBP reduction was greater in older than in younger subgroups. In Yin et al. 2022, pooled reduction estimates were −5.64 mmHg (95 % CI −7.39 to −3.89) for age ≥60 years and −3.47 mmHg (95 % CI −5.59 to −1.35) for age <60 years. In Tsai et al. 2022, the corresponding estimates were −5.95 mmHg (95 % CI −7.43 to −4.46) and −2.57 mmHg (95 % CI −4.13 to −1.00). In Greenwood et al. 2024, they were −7.15 mmHg (95 % CI −9.56 to −4.73) and −2.97 mmHg (95 % CI −4.52 to −1.41). Jafarnejad et al. 2020 used a 65-year cutoff and reported similarly large reductions in both age strata: −9.98 mmHg (95 % CI −14.06 to −5.90) for age ≥65 years and −10.38 mmHg (95 % CI −16.16 to −4.60) for age <65 years.

Baseline blood-pressure subgroup estimates were available in three reviews (**Supplementary Figure S2**). In hypertensive subgroups, pooled SBP estimates were −5.92 mmHg (95 % CI −7.81 to −4.03) in Yin et al. 2022, −5.07 mmHg (95 % CI −6.70 to −3.44) in Tsai et al. 2022, and −5.98 mmHg (95 % CI −8.88 to −3.08) in Greenwood et al. 2024. In the comparison subgroups with lower baseline blood pressure, estimates were −1.10 mmHg (95 % CI −12.83 to 10.63) in Yin et al. 2022, −5.39 mmHg (95 % CI −8.01 to −2.77) in Tsai et al. 2022, and −3.90 mmHg (95 % CI −5.27 to −2.53) in Greenwood et al. 2024.

### Formulation heterogeneity and network evidence

Reported potassium-containing LSSS formulations were heterogeneous across the included reviews (**Supplementary Table S6**). When explicitly stated, KCl content generally ranged from 25% to 66%, although published formulation classifications in Lai et al. 2026 extended from 3% to 66% KCl and were not mutually exclusive. Formal formulation analyses were limited (**Supplementary Figure S3**). In Yin et al. 2022, meta regression showed that each 10% lower NaCl content in the substitute was associated with a larger reduction in SBP (MD −1.53 mmHg, 95% CI −3.02 to −0.03). Greenwood et al. 2024 reported a prespecified subgroup analysis in which trials using ≥30% KCl showed a larger pooled SBP reduction (MD −7.88 mmHg, 95% CI −9.45 to −6.30; k=4) than trials using <30% KCl (MD −3.51 mmHg, 95% CI −4.91 to −2.10; k=7; P for interaction <0.01). Lai et al. 2026, the only included review to compare published formulation categories within a network meta-analysis, reported the largest SBP reduction for the high-potassium category (50–65% KCl; MD −7.54 mmHg, 95% CI −11.63 to −3.44), followed by the moderate-potassium category (25–40% KCl; MD −4.64 mmHg, 95% CI −6.05 to −3.24), the low-sodium category (60–79% NaCl; MD −4.39 mmHg, 95% CI −6.02 to −2.75), and the broader K-salt category (MD −4.00 mmHg, 95% CI −5.67 to −2.32). Because the Lai et al. 2026 formulation categories overlapped and could include mixed formulations with additional constituents, these estimates were treated as published formulation classifications rather than independent protocol-specific pooled estimates.

## Discussion

This umbrella review indicates that potassium-containing LSSS were consistently associated with lower systolic and diastolic blood pressure across overlapping review literature in mixed adult populations. Later reviews also reported associations in the same favourable direction for all-cause mortality, cardiovascular mortality, composite cardiovascular events, and selected stroke outcomes. However, the clinical-outcome evidence is less independently replicated than the blood-pressure evidence because later reviews repeatedly drew on the same primary trials, particularly SSaSS.^[19]^ Nonetheless, these patterns are consistent with broader umbrella reviews of dietary sodium and cardiovascular health, which report lower cardiovascular risk with lower sodium exposure and support sodium reduction or sodium substitution as a plausible dietary strategy for cardiovascular prevention.^[6,48–51]^

Blood pressure provides the most consistent Tier 2 signal in this umbrella review, with pooled SBP estimates ranging from −4.61 to −8.87 mmHg and pooled DBP estimates from −1.42 to −4.04 mmHg across the included reviews, all point estimates favouring potassium-containing LSSS. The clinical meaning of these reductions depends on baseline SBP, absolute cardiovascular risk, duration of exposure, and sustainability of the response, factors that statistical significance alone cannot capture. Within the causal framework linking urinary biomarker findings to Tier 1 clinical outcomes, SBP therefore serves as the most informative intermediate outcome.^[6,52,53]^ A 3–5 mmHg reduction may be important at the population level and particularly consequential for individuals near a diagnostic or treatment threshold, but the same absolute change carries different clinical weight across baseline-risk groups.^[54]^ Available subgroup data by hypertension status support this caution: estimates from hypertensive subgroups were consistently favourable, whereas data from lower-baseline-BP subgroups were fewer, more heterogeneously defined, and less consistent — suggesting, but not proving, that baseline BP modifies the clinical implications of potassium-containing LSSS. This pattern is consistent with the broader sodium-reduction literature, in which larger BP reductions are observed in hypertensive individuals and in chronic kidney disease populations.^[55–57]^

Urinary sodium and potassium excretion help clarify whether potassium-containing LSSS achieved the intended electrolyte shift. Across directly comparable reviews, pooled urinary sodium estimates were lower and pooled urinary potassium estimates were higher with potassium-containing LSSS. Tsai et al. (2022) was the only included review to report a pooled 24-h urinary Na:K ratio, and this remains a gap worth addressing. The Na:K ratio captures the combined directional shift in both electrolytes more directly than either analyte alone, and prospective data demonstrate that higher sodium excretion, lower potassium excretion, and an elevated Na:K ratio are each independently associated with greater cardiovascular risk.^[49,58]^ More consistent reporting of all three biomarkers (e.g., 24-h urinary sodium, 24-h urinary potassium, and urinary Na:K ratio) would therefore strengthen mechanistic interpretation in future potassium-containing LSSS trials and evidence syntheses.^[13,21]^

The Tier 1 clinical outcome evidence was directionally favourable but less uniform than the blood-pressure findings. All-cause and cardiovascular mortality estimates clustered below 1.00 across the later reviews, indicating a consistent direction of effect toward reduced risk. Major cardiovascular event estimates were more variable, largely reflecting differences in outcome definitions: Yin et al. (2022) and Greenwood et al. (2024) applied standard MACE-style definitions, whereas Brand et al. (2022) reported a broader, non-standardised cardiovascular composite that is not directly comparable. Future trials and evidence syntheses should therefore prioritise pre-specified, standardised cardiovascular composite definitions and report individual component outcomes separately.^[59]^ The available clinical-outcome literature is directionally consistent with SSaSS and with broader umbrella reviews linking lower sodium exposure to more favourable cardiovascular outcomes, but remains less independently replicated than the blood-pressure evidence.^[19,48,49]^

The registered protocol planned to distinguish primary- and secondary-prevention populations, but this distinction could not be operationalised reliably from the available review literature. Most included reviews classified populations by age, blood-pressure status, geography, or broad cardiovascular risk rather than by established CVD history, and the contributing RCTs were generally not designed to estimate effects separately among adults with versus without prior cardiovascular events. Future trials and evidence syntheses should pre-specify CVD history at baseline and report outcome estimates separately for participants with and without prior cardiovascular events.

The corrected covered area indicated very high primary-study overlap across the included reviews^[26]^, meaning that concordant review-level estimates reflect consistency rather than independent replication. The concentration of overlap is most pronounced around SSaSS, which anchors much of the later clinical-endpoint evidence: in Nguyen (2026), SSaSS is the sole trial contributing to the stroke outcome comparison for potassium-containing LSSS in the network, meaning its result entirely determines that estimate. SSaSS therefore provides the strongest direct evidence currently available for associations between potassium-containing LSSS and hard clinical outcomes, but within a specific context: rural Chinese populations with high discretionary salt use and substantial vascular risk burden. Additional large, randomised trials in populations outside East Asia, and in settings where processed foods rather than discretionary salt predominate as sodium sources, are needed to assess whether the observed clinical associations generalise across dietary contexts and baseline cardiovascular risk profiles.^[13,19,23,24]^

Formulation heterogeneity is a central feature of this evidence base. Across the included reviews, explicitly reported potassium-containing LSSS formulations spanned approximately 25% to 65% KCl, and many also included magnesium salts, calcium salts, folate, potassium citrate, lysine, flavorings, or other constituents.^[60,61]^ The NaCl:KCl composition determines the degree of sodium replacement and potassium exposure, while additional constituents may affect palatability and comparability across trials, both of which are relevant to interpretation. Taste-testing evidence from South Africa further illustrates this: a 50/50 KCl/NaCl blend was better accepted than higher-KCl formulations, highlighting that efficacy, acceptability, and scalability will need to be considered together in future formulation work.^[62]^ Lai et al. (2026) was the only included review to compare formulation categories directly within a network meta-analysis, but these categories were not mutually exclusive and did not map cleanly onto protocol-specific NaCl–KCl compositions. Confidence intervals overlapped substantially across potassium-content categories, and no clear dose-response gradient by KCl percentage was apparent. Greenwood et al. (2024) similarly found no monotonic relationship between KCl percentage and serum potassium in subgroup analyses. Together, these findings indicate that formulation effects cannot be inferred from KCl percentage alone, and that future primary trials should report full composition, co-constituents, iodization status, and delivery context in a standardised way.^[23,24,61]^

The safety evidence is more limited in scope than the efficacy evidence. Fewer reviews reported pooled safety outcomes, and those available were concentrated in serum potassium, hyperkalaemia events, and composite serious adverse events. Pooled serum potassium estimates ranged from −0.02 to +0.18 mmol/L, with four of five point estimates above zero. Brand et al. (2022) reported RR 1.04 (95% CI 0.46–2.38) for hyperkalaemia events, and Greenwood et al. (2024) reported RR 1.04 (95% CI 0.87–1.25) for serious adverse events. These estimates do not indicate a clear excess signal, but they apply primarily to selected populations rather than to those at highest risk of impaired potassium handling. Many contributing trials excluded participants with advanced chronic kidney disease, impaired potassium excretion, or use of potassium-sparing medications. The WHO 2025 LSSS guideline formalises this distinction, recommending potassium-containing LSSS for adults in general populations while explicitly excluding those with kidney impairment or other conditions that may compromise potassium excretion.^[15]^ For the remainder of the adult population, broader assessments conclude that benefits are likely to outweigh risks, but explicit contraindication messaging remains essential for groups with impaired potassium excretion.^[13,60]^ Future trials and post-implementation studies should directly examine potassium-containing LSSS in people with chronic kidney disease, reduced eGFR, RAAS blockade, or potassium-sparing diuretic use — groups that remain underrepresented in the randomised evidence but are central to safety guidance.^[15,63,64]^

Policy relevance now depends not only on efficacy but on whether potassium-containing LSSS can be introduced in real food environments with appropriate safety messaging. Although implementation outcomes were sparse in the included reviews, the broader literature indicates that consumer awareness, taste, price, labelling, iodisation compatibility, retail availability, and clear safety communication for groups in whom potassium-containing LSSS may be inappropriate are all likely to shape uptake.^[13,15,18,65]^ Formulation heterogeneity matters in this context because products differing in KCl content and co-constituents may differ in taste, acceptability, and clarity of contraindication messaging. Recent reviews in primary care and public health increasingly describe sodium substitution as a pragmatic dietary strategy, emphasising that it must be implemented within real purchasing and cooking environments rather than treated solely as a trial intervention.^[15,23]^. A narrative review of systematic reviews in primary care positioned sodium reduction and substitution among the more practical dietary strategies for cardiovascular risk management when adapted to routine food purchasing, cooking, and counselling contexts.^[66]^

This umbrella review has several strengths and limitations. Strengths include prospective registration, duplicate screening and extraction, AMSTAR 2 quality appraisal, and formal quantification of primary-study overlap — design features that are particularly important in a literature where the same core trials recur across multiple later reviews. The main limitations arise from the source evidence. Prevention status was often mixed or unclear at review level, formulation reporting was inconsistent, and the clinical-endpoint literature was heavily influenced by SSaSS. A further limitation is that pooled estimates from three otherwise eligible reviews could not be fully disentangled from two non-potassium-chloride chitosan interventions.^[67,68]^ Those affected summary estimates were retained as reported but treated as partially extractable, which limits precision in protocol-specific effect-size estimation for potassium-containing LSSS.

Several research priorities emerge from these findings. First, clinical-outcome evidence remains geographically concentrated, and additional trials outside East Asian settings are needed. Second, formulation-specific comparisons require greater emphasis. Available subgroup analyses suggested that SBP associations differed across published formulation strata, yet the field continues to pool heterogeneous products under a single ‘salt substitute’ label. Future primary trials and evidence syntheses should report and compare mutually exclusive formulation categories. Third, future trials should consistently report serum potassium, kidney outcomes, and urinary Na:K ratio, particularly in participants using Renin-Angiotensin-Aldosterone System inhibitors or those at potential kidney-related risk. Fourth, implementation research should extend beyond efficacy to address affordability, labelling, and food-system integration. Notably, ongoing and recently registered trials are beginning to address some of these gaps. A trial registered in Düsseldorf is evaluating the vascular and immune effects of potassium chloride substitution (ClinicalTrials.gov: NCT05970601), and a family-based South African protocol is evaluating a potassium-enriched LSSS for urinary Na:K ratio and blood pressure in adolescents and their households [PACTR202306727520808].^[62]^

## Conclusion

Potassium-containing LSSS were consistently associated with lower blood pressure across the included review literature, and the available clinical-endpoint evidence was directionally aligned with lower fatal stroke, haemorrhagic stroke, all-cause mortality and cardiovascular mortality. The main remaining uncertainties concern formulation, generalisability beyond the settings that dominate the current evidence base, differentiation by baseline CVD history, and safety in populations with impaired potassium handling. These findings support potassium-containing LSSS as a promising nutritional strategy for cardiovascular prevention in mixed adult populations, while underscoring the need for more precise trial reporting and implementation-specific safety guidance.

## Data Availability

All data produced in the present study are available upon reasonable request to the authors

## Acknowledgements

The authors thank William R. Garcia for assistance with translation of a Portuguese-language full-text report during full-text screening, Miyu Niwa for comments on manuscript clarity, and Ziwei He and Dr. Ensheng Dong for assistance with the R code used to generate the forest plots.

## List of abbreviations

AKI: acute kidney injur
AMSTAR 2: A Measurement Tool to Assess Systematic Reviews 2
BP: blood pressure
CCA: corrected covered area
CKD: chronic kidney disease
CVD: cardiovascular disease
DBP: diastolic blood pressure
eGFR: estimated glomerular filtration rate
HR: hazard ratio
KCl: potassium chloride
LSSS: low-sodium salt substitute
MACE: major adverse cardiovascular events (used only where reported by source reviews)
Na:K: sodium-to-potassium ratio
NaCl: sodium chloride
NMA: network meta-analysis
RCT: randomised controlled trial
RR: risk ratio or rate ratio as reported by source reviews
SAE: serious adverse event
SBP: systolic blood pressure
SR: systematic review
SSaSS: Salt Substitute and Stroke Study.

