## Supplementary File for "Efficacy and safety of potassium-containing low-sodium salt substitutes for cardiovascular disease prevention in mixed adult populations: an umbrella review"

**Supplementary Figure S1. Pairwise overlap heatmap across included reviews.**

|  | **Hernandez 2019** |  |  |  |  |  |  |  |  |  |
| --- | --- | --- | --- | --- | --- | --- | --- | --- | --- | --- |
|  |  | **Jafarnejad 2020** | **Jin 2020** | **Yin 2022** | **Brand 2022** | **Tsai 2022** | **Aliasgharzadeh 2022** | **Greenwood 2024** | **Prommas 2026** |  |
| **Jafarnejad 2020** | 52.6% |  |  |  |  |  |  |  |  |  |
| **Jin 2020** | 26.1% | 17.6% |  |  |  |  |  |  |  |  |
| **Yin 2022** | 56.5% | 35.0% | 28.6% |  |  |  |  |  |  |  |
| **Brand 2022** | 42.9% | 24.0% | 24.0% | 52.0% |  |  |  |  |  |  |
| **Tsai 2022** | 35.7% | 20.8% | 20.8% | 56.5% | 48.1% |  |  |  |  | **Nguyen 2026** |
| **Aliasgharzadeh 2022** | 30.4% | 23.5% | 50.0% | 47.4% | 33.3% | 42.9% |  |  |  |  |
| **Greenwood 2024** | 8.3% | 0.0% | 13.3% | 20.0% | 16.7% | 30.0% | 28.6% |  |  |  |
| **Prommas 2026** | 9.1% | 15.4% | 25.0% | 22.2% | 18.2% | 26.3% | 33.3% | 20.0% |  |  |
| **Nguyen 2026** | 0.0% | 0.0% | 0.0% | 0.0% | 4.8% | 5.3% | 9.1% | 14.3% | 20.0% |  |
| **Lai 2026** | 55.9% | 29.4% | 22.2% | 45.7% | 57.1% | 39.5% | 28.6% | 17.1% | 11.4% | 2.9% |

Heatmap showing pairwise overlap in primary studies between included, protocol-eligible reviews. Each cell represents the pairwise corrected covered area (CCA) for a given review pair, with darker shading indicating greater overlap. Pairwise overlap was classified as slight (<5 %), moderate (5 to <10 %), high (10 to <15 %), or very high (≥15 %). Abbreviation: CCA, corrected covered area.

**Supplementary Figure S2. Review-level subgroup analyses of systolic blood pressure associated with potassium-containing low-sodium salt substitutes.**

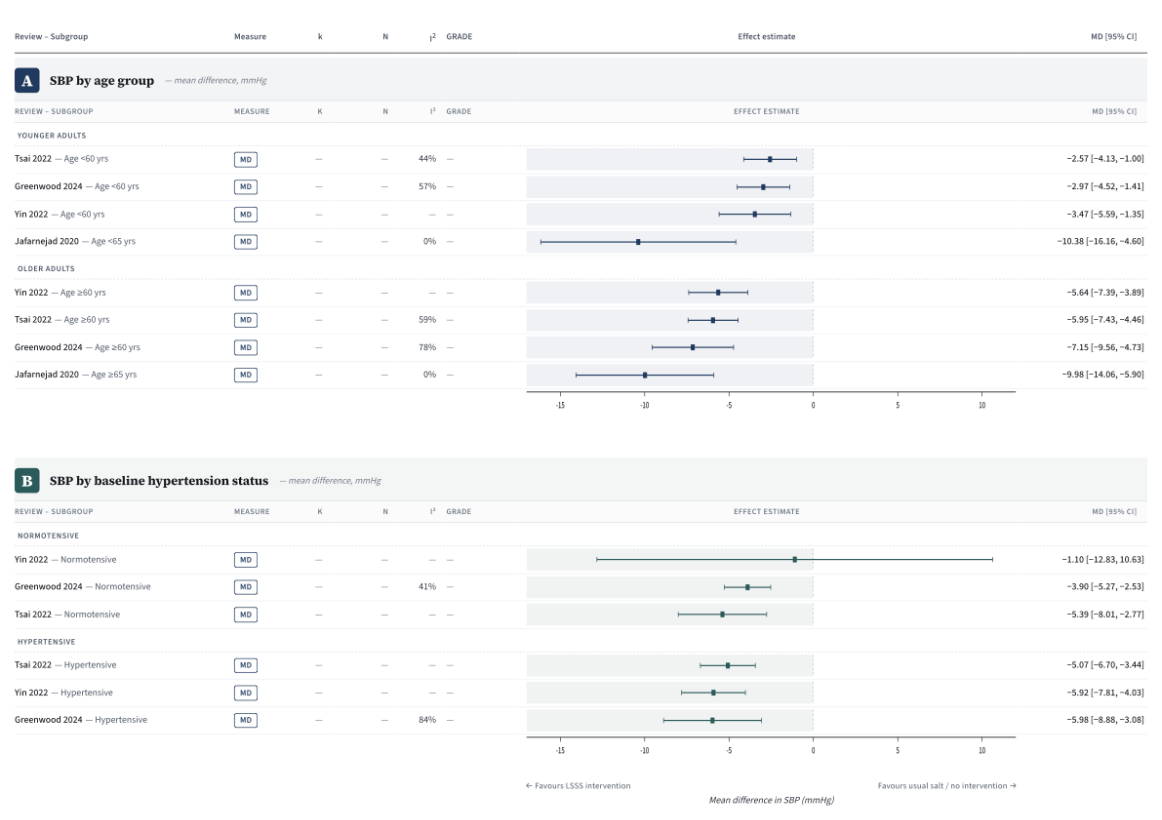

Panels show pooled subgroup estimates for systolic blood pressure by age category and by baseline hypertension status, as reported by the source reviews. Age subgroup thresholds followed the source reviews (≥60 v. <60 years in Yin 2022, Tsai 2022 and Greenwood 2024; ≥65 v. <65 years in Jafarnejad 2020). Hypertension subgroup categories were reproduced as reported by the source reviews. Estimates were not pooled across reviews.

**Supplementary Figure S3. Systolic blood pressure by published potassium chloride content or formulation category.**

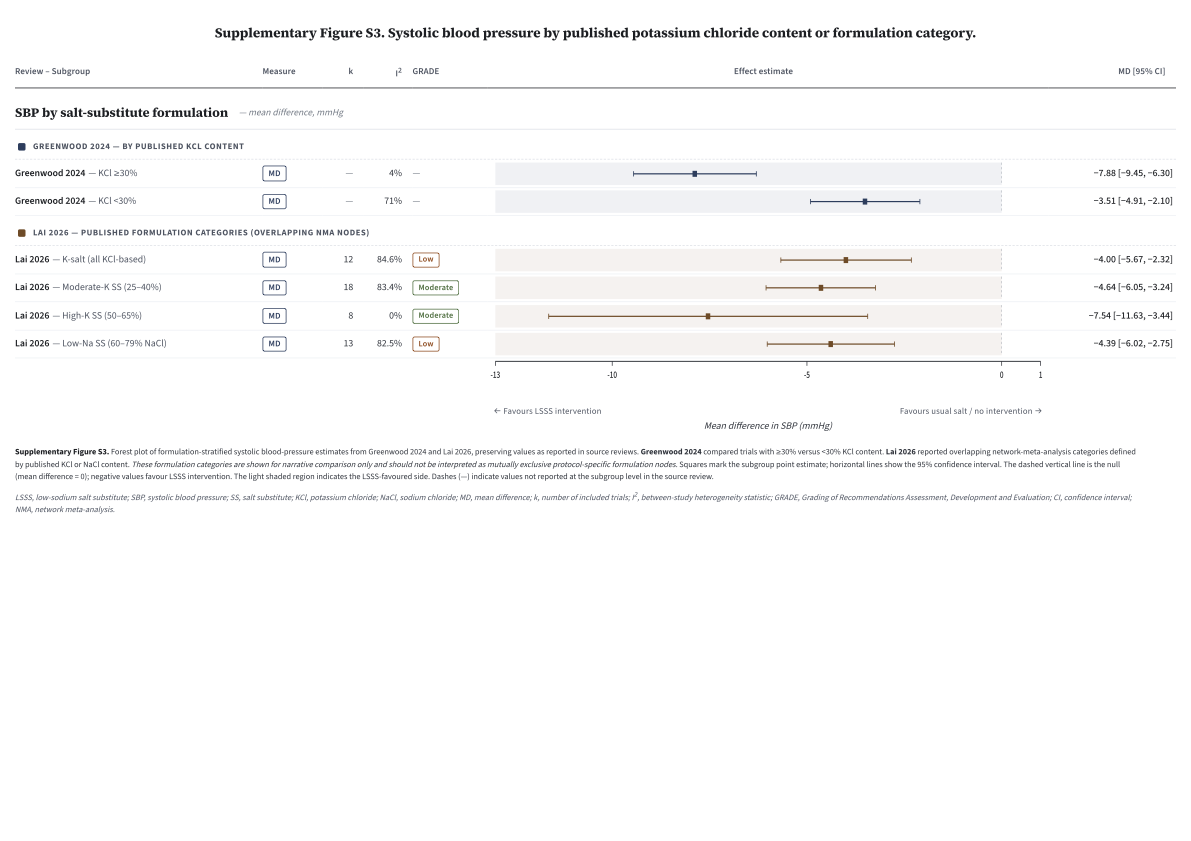

Forest plot showing published formulation-stratified systolic blood-pressure estimates from Greenwood 2024 and Lai 2026. Greenwood 2024 compared trials with ≥30 % v. <30 % KCl. Lai 2026 reported overlapping network-meta-analysis categories based on published KCl or NaCl content. These formulation categories are shown for narrative comparison only and should not be interpreted as mutually exclusive protocol-specific formulation nodes.

**Supplementary Table S1. Eligibility criteria and justification of excluded record/study types for the umbrella review.**

**Panel A. Eligibility criteria and rationale**

| **Domain** | **Included** | **Excluded** | **Rationale** |
| --- | --- | --- | --- |
| **Review type** | Systematic reviews with or without meta-analysis, meta-analyses, and umbrella reviews reporting a reproducible search and study-selection process. | Narrative or non-systematic reviews, editorials, commentaries, protocols/registrations, conference abstracts without sufficient review data, and single primary studies. | The umbrella review was designed to synthesise review-level evidence rather than individual primary studies, and only reviews with reproducible methods could support valid comparison of overlapping evidence. |
| **Population** | Primary- and/or secondary-prevention populations of relevance to cardiovascular disease prevention. | Reviews focused on conditions outside the cardiovascular prevention scope. | The review question addressed potassium-containing low-sodium salt substitutes in relation to blood pressure, cardiovascular events, mortality, validated intake biomarkers, and safety outcomes relevant to cardiovascular prevention. |
| **Intervention** | Potassium-containing low-sodium salt substitutes used in cooking, seasoning, food preparation, household/community distribution, or food reformulation when extractable. | Potassium supplements in pill/capsule form; non-potassium salt alternatives (e.g. monosodium glutamate, herb blends); general sodium-reduction advice or dietary patterns without an extractable potassium-containing salt-substitute component. | The intervention of interest required partial replacement of sodium chloride with potassium-containing salt substitute, most commonly potassium chloride, rather than general sodium reduction alone or non-potassium salt alternatives. |
| **Comparator** | Regular salt, usual care, placebo, no intervention, or another comparator permitting extraction of the potassium-containing salt-substitute effect. | Comparators that did not allow separation of the potassium-containing salt-substitute effect from co-interventions. | Comparators needed to preserve interpretability of the potassium-containing salt-substitute contrast. |
| **Outcomes** | Review-level reporting of at least one eligible efficacy or safety outcome, including Tier 1 clinical outcomes, Tier 2 biomarkers/intermediate outcomes, Tier 3 safety outcomes, or implementation-relevant outcomes. | Reports without extractable outcome data relevant to the umbrella review question. | At least one relevant outcome was required to support evidence synthesis and evidence mapping. |
| **Language and date** | No language or publication-date restrictions were applied at the umbrella-review level. | None a priori; studies published after the final search date were not eligible. | This approach maximised sensitivity while preserving consistency with the registered protocol and fixed search cut-off. |

**Panel B. Full-text exclusion categories**

| **Full-text exclusion category** | **n** | **Operational definition** | **Reason for exclusion** |
| --- | --- | --- | --- |
| Wrong intervention (not potassium-containing LSSS or not extractable) | 35 | Review did not assess potassium-containing low-sodium salt substitutes, or LSSS-specific data could not be separated from broader interventions. | Records did not address the target intervention or did not permit extraction of potassium-containing LSSS results. |
| Not an eligible review type | 14 | Publication was not a systematic review, meta-analysis, or umbrella review with reproducible methods. | Non-eligible review designs could not support review-level evidence synthesis. |
| Abstract-only publication | 12 | Conference abstract, oral session abstract, or similar short-form record without sufficient methodological detail or extractable review data. | Insufficient information was available to assess eligibility, methods, or extract outcomes reliably. |
| Wrong intervention and wrong condition/domain | 8 | Record was outside both the intervention scope and cardiovascular prevention scope. | These records did not contribute evidence relevant to the review question. |
| Editorial | 1 | Editorial or opinion piece without systematic review methods. | Did not provide review-level evidence. |
| Protocol/registration only | 1 | Protocol or registration record without completed review results. | No results were available for synthesis. |
| Wrong condition/domain | 1 | Record addressed a health domain outside cardiovascular prevention. | Outside the scope of the umbrella review question. |
| **Total full-text exclusions** | **72** |  |  |

Panel A summarises the prespecified eligibility criteria by domain. Panel B summarises the full-text exclusion categories and counts corresponding to Figure 2. Full-text exclusion counts correspond to the PRISMA 2020 flow diagram in Figure 2. This supplementary table provides category-level justification for excluded record/study types. If a citation-level list of all excluded full-text reports is required for peer review, it can be generated from the Covidence full-text exclusion export.

**Supplementary Table S2. Final electronic search strategies and retrieval counts.**

Database-specific search strategies used for PubMed, Embase, Web of Science, Global Health (EBSCO) and the Cochrane Database of Systematic Reviews, including final search dates and numbers of records retrieved.

| **Database** | **Search String** | **Date** | **Results** |
| --- | --- | --- | --- |
| [PubMed](https://www.offcampus.lib.washington.edu/login?url=https://pubmed.ncbi.nlm.nih.gov/?myncbishare=uwonline) | ( ("Potassium, dietary"[Mesh] OR "potassium dietary"[Supplementary Concept] OR "potassium chloride"[Supplementary Concept] OR "potassium chloride"[Mesh] OR "potassium chloride"[tiab] OR "potassium citrate"[Supplementary Concept] OR "potassium citrate"[Mesh] OR "potassium citrate"[tiab] OR "potassium salt"[tiab:~2] OR "KCl salt"[tiab] OR "salt substitut*"[tiab] OR "salt substitute"[tiab:~2] OR "salt substitution"[tiab:~2] OR "sodium substitut*"[tiab] OR "sodium substitute"[tiab:~2] OR "potassium enrich*"[tiab] OR "potassium rich*"[tiab] OR "reduce* sodium salt"[tiab] OR "low sodium salt"[tiab:~2] OR "salt reduction"[tiab:~2] OR "salt replac*"[tiab] OR "reduc* salt content"[tiab] OR "reduc* sodium content"[tiab] OR "reduc* dietary sodium"[tiab] OR "reduc* dietary salt"[tiab] OR "reduc* salt intake"[tiab] OR "reduc* sodium intake"[tiab] OR "low salt"[tiab:~2] OR "low sodium"[tiab:~2] OR "sodium free"[tiab:~2] OR "discretionary salt"[tiab:~2]) AND ("Dietary Approaches To Stop Hypertension"[Mesh] OR "Hypertension"[Mesh] OR "Blood Pressure"[Mesh] OR "Cardiovascular Diseases"[Mesh] OR "Heart Diseases"[Mesh] OR "Heart Failure"[Mesh] OR "Myocardial Ischemia"[Mesh] OR "Stroke"[Mesh] OR "Ischemic Stroke"[Mesh] OR "Intracranial Hemorrhages"[Mesh] OR "Cerebral Hemorrhage"[Mesh] OR "Subarachnoid Hemorrhage"[Mesh] OR normotens*[tiab] OR hypertens*[tiab] OR "blood pressure"[tiab] OR "cardiovascular disease*"[tiab] OR "heart disease*"[tiab] OR "heart failure"[tiab] OR "myocardial infarct*"[tiab] OR "heart attack*"[tiab] OR "ischemic heart"[tiab] OR "ischaemic heart"[tiab] OR stroke[tiab] OR "intracerebral hemorrhag*"[tiab] OR "intracerebral haemorrhag*"[tiab] OR "cerebral hemorrhag*"[tiab] OR "cerebral haemorrhag*"[tiab] OR "intracranial hemorrhag*"[tiab] OR "intracranial haemorrhag*"[tiab] OR "cranial hemorrhag*"[tiab] OR "cranial haemorrhag*"[tiab] OR "subarachnoid hemorrhag*"[tiab] OR "subarachnoid haemorrhag*"[tiab] OR "brain infarct*"[tiab] OR "cerebral infarct*"[tiab] OR "cerebrovascular accident*"[tiab] OR "death"[Mesh] OR "death"[tiab] ) ) AND (("meta analysis"[Publication Type] OR "systematic review"[Publication Type] OR meta analysis as topic[MeSH Terms] OR "systematic reviews as topic"[MeSH Terms] OR systematic[sb] OR "systematic review"[tiab:~2] OR "systematically reviewed"[tiab:~2] OR "meta analysis" OR "meta analyses" OR metaanaly* OR "cochrane review" OR "critical review" OR "evidence gap map" OR "integrative review" OR "mapping review" OR "mixed studies review" OR "mixed methods review" OR "evidence synthesis" OR "research synthesis" OR "rapid review" OR "realist review" OR "scoping review"[tiab:~2] OR "state-of-the-art review" OR "systematized review" OR "umbrella review" OR "review of reviews" OR "The Cochrane database of systematic reviews"[Journal])) | 03/06/2026 | 332 (4 ) |
| [Embase](https://www.offcampus.lib.washington.edu/login?url=https://www.embase.com/#advancedSearch/default) | ('potassium intake'/exp OR 'potassium intake' OR 'potassium dietary'/exp OR 'potassium dietary' OR 'potassium chloride'/exp OR 'potassium chloride' OR 'potassium chloride':ti,ab,kw OR 'potassium citrate'/exp OR 'potassium citrate' OR 'citrate potassium'/exp OR 'citrate potassium' OR 'potassium citrate':ti,ab,kw OR (('potassium' NEAR/3 'salt'):ti,ab,kw) OR 'kcl salt':ti,ab,kw OR 'salt substitut*':ti,ab,kw OR (('salt' NEAR/3 'substitute'):ti,ab,kw) OR (('salt' NEAR/3 'substitution'):ti,ab,kw) OR 'sodium substitut*':ti,ab,kw OR (('sodium' NEAR/3 'substitute'):ti,ab,kw) OR 'potassium enrich*':ti,ab,kw OR 'potassium rich*':ti,ab,kw OR 'reduce* sodium salt':ti,ab,kw OR (('low' NEAR/3 'sodium' NEAR/3 'salt'):ti,ab,kw) OR (('salt' NEAR/3 'reduction'):ti,ab,kw) OR 'salt replac*':ti,ab,kw OR 'reduc* salt content':ti,ab,kw OR 'reduc* sodium content':ti,ab,kw OR 'reduc* dietary sodium':ti,ab,kw OR 'reduc* dietary salt':ti,ab,kw OR 'reduc* salt intake':ti,ab,kw OR 'reduc* sodium intake':ti,ab,kw OR (('low' NEAR/3 'salt'):ti,ab,kw) OR (('low' NEAR/3 'sodium'):ti,ab,kw) OR (('sodium' NEAR/3 'free'):ti,ab,kw) OR (('discretionary' NEAR/3 'salt'):ti,ab,kw)) AND ('dash diet'/exp OR 'dash diet' OR 'hypertension'/exp OR 'hypertension' OR 'blood pressure'/exp OR 'blood pressure' OR 'cardiovascular disease'/exp OR 'cardiovascular disease' OR 'heart disease'/exp OR 'heart disease' OR 'heart failure'/exp OR 'heart failure' OR 'heart muscle ischemia'/exp OR 'heart muscle ischemia' OR 'cerebrovascular accident'/exp OR 'cerebrovascular accident' OR 'ischemic stroke'/exp OR 'ischemic stroke' OR 'brain hemorrhage'/exp OR 'brain hemorrhage' OR 'subarachnoid hemorrhage'/exp OR 'subarachnoid hemorrhage' OR 'normotens*':ti,ab,kw OR 'hypertens*':ti,ab,kw OR 'blood pressure':ti,ab,kw OR 'cardiovascular disease*':ti,ab,kw OR 'heart disease*':ti,ab,kw OR 'heart failure':ti,ab,kw OR 'myocardial infarct*':ti,ab,kw OR 'heart attack*':ti,ab,kw OR 'ischemic heart':ti,ab,kw OR 'ischaemic heart':ti,ab,kw OR 'stroke':ti,ab,kw OR 'intracerebral hemorrhag*':ti,ab,kw OR 'intracerebral haemorrhag*':ti,ab,kw OR 'cerebral hemorrhag*':ti,ab,kw OR 'cerebral haemorrhag*':ti,ab,kw OR 'intracranial hemorrhag*':ti,ab,kw OR 'intracranial haemorrhag*':ti,ab,kw OR 'cranial hemorrhag*':ti,ab,kw OR 'cranial haemorrhag*':ti,ab,kw OR 'subarachnoid hemorrhag*':ti,ab,kw OR 'subarachnoid haemorrhag*':ti,ab,kw OR 'brain infarct*':ti,ab,kw OR 'cerebral infarct*':ti,ab,kw OR 'cerebrovascular accident*':ti,ab,kw OR 'death'/exp OR 'death' OR 'death':ti,ab,kw) AND ('meta analysis'/de OR 'systematic review'/de OR 'meta analysis topic'/de OR 'systematic review topic'/de) AND [embase]/lim NOT ([embase]/lim AND [medline]/lim) | 03/06/2026 | 232 (4) |
| [Global Health (EBSCO)](https://offcampus.lib.washington.edu/login?url=https://search.ebscohost.com/login.aspx?authtype=ip,uid&profile=ehost&defaultdb=lhh) | (((MH "Potassium, dietary+") OR (MW "potassium dietary") OR (MW "potassium chloride") OR (MH "potassium chloride+") OR (TI "potassium chloride" OR AB "potassium chloride") OR (MW "potassium citrate") OR (MH "potassium citrate+") OR (TI "potassium citrate" OR AB "potassium citrate") OR "potassium salt[tiab:~2]" OR (TI "KCl salt" OR AB "KCl salt") OR (TI "salt substitut*" OR AB "salt substitut*") OR "salt substitute[tiab:~2]" OR "salt substitution[tiab:~2]" OR (TI "sodium substitut*" OR AB "sodium substitut*") OR "sodium substitute[tiab:~2]" OR (TI "potassium enrich*" OR AB "potassium enrich*") OR (TI "potassium rich*" OR AB "potassium rich*") OR (TI "reduce* sodium salt" OR AB "reduce* sodium salt") OR "low sodium salt[tiab:~2]" OR "salt reduction[tiab:~2]" OR (TI "salt replac*" OR AB "salt replac*") OR (TI "reduc* salt content" OR AB "reduc* salt content") OR (TI "reduc* sodium content" OR AB "reduc* sodium content") OR (TI "reduc* dietary sodium" OR AB "reduc* dietary sodium") OR (TI "reduc* dietary salt" OR AB "reduc* dietary salt") OR (TI "reduc* salt intake" OR AB "reduc* salt intake") OR (TI "reduc* sodium intake" OR AB "reduc* sodium intake") OR "low salt[tiab:~2]" OR "low sodium[tiab:~2]" OR "sodium free[tiab:~2]" OR "discretionary salt[tiab:~2]" ) AND ((MH "Dietary Approaches To Stop Hypertension+") OR (MH Hypertension+) OR (MH "Blood Pressure+") OR (MH "Cardiovascular Diseases+") OR (MH "Heart Diseases+") OR (MH "Heart Failure+") OR (MH "Myocardial Ischemia+") OR (MH Stroke+) OR (MH "Ischemic Stroke+") OR (MH "Intracranial Hemorrhages+") OR (MH "Cerebral Hemorrhage+") OR (MH "Subarachnoid Hemorrhage+") OR (TI normotens* OR AB normotens*) OR (TI hypertens* OR AB hypertens*) OR (TI "blood pressure" OR AB "blood pressure") OR (TI "cardiovascular disease*" OR AB "cardiovascular disease*") OR (TI "heart disease*" OR AB "heart disease*") OR (TI "heart failure" OR AB "heart failure") OR (TI "myocardial infarct*" OR AB "myocardial infarct*") OR (TI "heart attack*" OR AB "heart attack*") OR (TI "ischemic heart" OR AB "ischemic heart") OR (TI "ischaemic heart" OR AB "ischaemic heart") OR (TI stroke OR AB stroke) OR (TI "intracerebral hemorrhag*" OR AB "intracerebral hemorrhag*") OR (TI "intracerebral haemorrhag*" OR AB "intracerebral haemorrhag*") OR (TI "cerebral hemorrhag*" OR AB "cerebral hemorrhag*") OR (TI "cerebral haemorrhag*" OR AB "cerebral haemorrhag*") OR (TI "intracranial hemorrhag*" OR AB "intracranial hemorrhag*") OR (TI "intracranial haemorrhag*" OR AB "intracranial haemorrhag*") OR (TI "cranial hemorrhag*" OR AB "cranial hemorrhag*") OR (TI "cranial haemorrhag*" OR AB "cranial haemorrhag*") OR (TI "subarachnoid hemorrhag*" OR AB "subarachnoid hemorrhag*") OR (TI "subarachnoid haemorrhag*" OR AB "subarachnoid haemorrhag*") OR (TI "brain infarct*" OR AB "brain infarct*") OR (TI "cerebral infarct*" OR AB "cerebral infarct*") OR (TI "cerebrovascular accident*" OR AB "cerebrovascular accident*") OR (MH death+) OR (TI death OR AB death))) AND (((PT "meta analysis") OR (PT "systematic review") OR (MH "meta analysis as topic+") OR (MH "systematic reviews as topic+") OR (SB systematic) OR "systematic review[tiab:~2]" OR "systematically reviewed[tiab:~2]" OR "meta analysis" OR "meta analyses" OR metaanaly* OR "cochrane review" OR "critical review" OR "evidence gap map" OR "integrative review" OR "mapping review" OR "mixed studies review" OR "mixed methods review" OR "evidence synthesis" OR "research synthesis" OR "rapid review" OR "realist review" OR "scoping review[tiab:~2]" OR "state-of-the-art review" OR "systematized review" OR "umbrella review" OR "review of reviews" OR (SO "The Cochrane database of systematic reviews" OR ST "The Cochrane database of systematic reviews" OR IB "The Cochrane database of systematic reviews"))) | 03/06/2026 | 45 (1) |
| [Web of Science](https://www.offcampus.lib.washington.edu/login?url=https://www.webofscience.com/wos/woscc/basic-search) | ((TS=(("potassium dietary" OR "potassium chloride" OR "potassium citrate" OR "potassium salt" OR "KCl salt" OR "salt substitut*" OR "salt substitute" OR "salt substitution" OR "sodium substitut*" OR "sodium substitute" OR "potassium enrich*" OR "potassium rich*" OR "reduce* sodium salt" OR "low sodium salt" OR "salt reduction" OR "salt replac*" OR "reduc* salt content" OR "reduc* sodium content" OR "reduc* dietary sodium" OR "reduc* dietary salt" OR "reduc* salt intake" OR "reduc* sodium intake" OR "low salt" OR "low sodium" OR "sodium free" OR "discretionary salt") )) AND TS=(("Dietary Approaches To Stop Hypertension" OR "Hypertension" OR "Blood Pressure" OR "Cardiovascular Disease" OR "Heart Disease" OR "Heart Failure" OR "Myocardial Ischemia" OR "Stroke" OR "Ischemic Stroke" OR "Intracranial Hemorrhage" OR "Cerebral Hemorrhage" OR "Subarachnoid Hemorrhage" OR normotens* OR hypertens* OR "myocardial infarct*" OR "heart attack*" OR "ischemic heart" OR "ischaemic heart" OR "intracerebral hemorrhag*" OR "intracerebral haemorrhag*" OR "cerebral hemorrhag*" OR "cerebral haemorrhag*" OR "intracranial hemorrhag*" OR "intracranial haemorrhag*" OR "cranial hemorrhag*" OR "cranial haemorrhag*" OR "subarachnoid hemorrhag*" OR "subarachnoid haemorrhag*" OR "brain infarct*" OR "cerebral infarct*" OR "cerebrovascular accident*" OR "death"))) AND TS=(("meta analysis" OR "systematic review" OR "systematically reviewed" OR "meta analysis" OR "meta analyses" OR metaanaly* OR "cochrane review" OR "critical review" OR "evidence gap map" OR "integrative review" OR "mapping review" OR "mixed studies review" OR "mixed methods review" OR "evidence synthesis" OR "research synthesis" OR "rapid review" OR "realist review" OR "scoping review" OR "state-of-the-art review" OR "systematized review" OR "umbrella review" OR "review of reviews")) | 03/06/2026 | 572; 209 after refined to Document Type: Review Articles  (575; 210) |
| [Cochrane](http://offcampus.lib.washington.edu/login?url=https://www.cochranelibrary.com/) Systematic Reviews | ("potassium dietary" OR "potassium chloride" OR "potassium citrate" OR "potassium salt" OR "KCl salt" OR "salt substitut*" OR "salt substitute" OR "salt substitution" OR "sodium substitut*" OR "sodium substitute" OR "potassium enrich*" OR "potassium rich*" OR "reduce* sodium salt" OR "low sodium salt" OR "salt reduction" OR "salt replac*" OR "reduc* salt content" OR "reduc* sodium content" OR "reduc* dietary sodium" OR "reduc* dietary salt" OR "reduc* salt intake" OR "reduc* sodium intake" OR "low salt" OR "low sodium" OR "sodium free" OR "discretionary salt") in Title Abstract Keyword AND ("Dietary Approaches To Stop Hypertension" OR "Hypertension" OR "Blood Pressure" OR "Cardiovascular Disease" OR "Heart Disease" OR "Heart Failure" OR "Myocardial Ischemia" OR "Stroke" OR "Ischemic Stroke" OR "Intracranial Hemorrhage" OR "Cerebral Hemorrhage" OR "Subarachnoid Hemorrhage" OR normotens* OR hypertens* OR "myocardial infarct*" OR "heart attack*" OR "ischemic heart" OR "ischaemic heart" OR "intracerebral hemorrhag*" OR "intracerebral haemorrhag*" OR "cerebral hemorrhag*" OR "cerebral haemorrhag*" OR "intracranial hemorrhag*" OR "intracranial haemorrhag*" OR "cranial hemorrhag*" OR "cranial haemorrhag*" OR "subarachnoid hemorrhag*" OR "subarachnoid haemorrhag*" OR "brain infarct*" OR "cerebral infarct*" OR "cerebrovascular accident*" OR "death") in Title Abstract Keyword - in Cochrane Reviews (Word variations have been searched) | 03/06/2026 | 20 (no change) |

February Searches

| **Database** | **Search String** | **Date** | **Results** |
| --- | --- | --- | --- |
| [PubMed](https://www.offcampus.lib.washington.edu/login?url=https://pubmed.ncbi.nlm.nih.gov/?myncbishare=uwonline) | ( ("Potassium, dietary"[Mesh] OR "potassium dietary"[Supplementary Concept] OR "potassium chloride"[Supplementary Concept] OR "potassium chloride"[Mesh] OR "potassium chloride"[tiab] OR "potassium citrate"[Supplementary Concept] OR "potassium citrate"[Mesh] OR "potassium citrate"[tiab] OR "potassium salt"[tiab:~2] OR "KCl salt"[tiab] OR "salt substitut*"[tiab] OR "salt substitute"[tiab:~2] OR "salt substitution"[tiab:~2] OR "sodium substitut*"[tiab] OR "sodium substitute"[tiab:~2] OR "potassium enrich*"[tiab] OR "potassium rich*"[tiab] OR "reduce* sodium salt"[tiab] OR "low sodium salt"[tiab:~2] OR "salt reduction"[tiab:~2] OR "salt replac*"[tiab] OR "reduc* salt content"[tiab] OR "reduc* sodium content"[tiab] OR "reduc* dietary sodium"[tiab] OR "reduc* dietary salt"[tiab] OR "reduc* salt intake"[tiab] OR "reduc* sodium intake"[tiab] OR "low salt"[tiab:~2] OR "low sodium"[tiab:~2] OR "sodium free"[tiab:~2] OR "discretionary salt"[tiab:~2]) AND ("Dietary Approaches To Stop Hypertension"[Mesh] OR "Hypertension"[Mesh] OR "Blood Pressure"[Mesh] OR "Cardiovascular Diseases"[Mesh] OR "Heart Diseases"[Mesh] OR "Heart Failure"[Mesh] OR "Myocardial Ischemia"[Mesh] OR "Stroke"[Mesh] OR "Ischemic Stroke"[Mesh] OR "Intracranial Hemorrhages"[Mesh] OR "Cerebral Hemorrhage"[Mesh] OR "Subarachnoid Hemorrhage"[Mesh] OR normotens*[tiab] OR hypertens*[tiab] OR "blood pressure"[tiab] OR "cardiovascular disease*"[tiab] OR "heart disease*"[tiab] OR "heart failure"[tiab] OR "myocardial infarct*"[tiab] OR "heart attack*"[tiab] OR "ischemic heart"[tiab] OR "ischaemic heart"[tiab] OR stroke[tiab] OR "intracerebral hemorrhag*"[tiab] OR "intracerebral haemorrhag*"[tiab] OR "cerebral hemorrhag*"[tiab] OR "cerebral haemorrhag*"[tiab] OR "intracranial hemorrhag*"[tiab] OR "intracranial haemorrhag*"[tiab] OR "cranial hemorrhag*"[tiab] OR "cranial haemorrhag*"[tiab] OR "subarachnoid hemorrhag*"[tiab] OR "subarachnoid haemorrhag*"[tiab] OR "brain infarct*"[tiab] OR "cerebral infarct*"[tiab] OR "cerebrovascular accident*"[tiab] OR "death"[Mesh] OR "death"[tiab] ) ) AND (("meta analysis"[Publication Type] OR "systematic review"[Publication Type] OR meta analysis as topic[MeSH Terms] OR "systematic reviews as topic"[MeSH Terms] OR systematic[sb] OR "systematic review"[tiab:~2] OR "systematically reviewed"[tiab:~2] OR "meta analysis" OR "meta analyses" OR metaanaly* OR "cochrane review" OR "critical review" OR "evidence gap map" OR "integrative review" OR "mapping review" OR "mixed studies review" OR "mixed methods review" OR "evidence synthesis" OR "research synthesis" OR "rapid review" OR "realist review" OR "scoping review"[tiab:~2] OR "state-of-the-art review" OR "systematized review" OR "umbrella review" OR "review of reviews" OR "The Cochrane database of systematic reviews"[Journal])) | 02/03/2026 | 328 |
| [Embase](https://www.offcampus.lib.washington.edu/login?url=https://www.embase.com/#advancedSearch/default) | ('potassium intake'/exp OR 'potassium intake' OR 'potassium dietary'/exp OR 'potassium dietary' OR 'potassium chloride'/exp OR 'potassium chloride' OR 'potassium chloride':ti,ab,kw OR 'potassium citrate'/exp OR 'potassium citrate' OR 'citrate potassium'/exp OR 'citrate potassium' OR 'potassium citrate':ti,ab,kw OR (('potassium' NEAR/3 'salt'):ti,ab,kw) OR 'kcl salt':ti,ab,kw OR 'salt substitut*':ti,ab,kw OR (('salt' NEAR/3 'substitute'):ti,ab,kw) OR (('salt' NEAR/3 'substitution'):ti,ab,kw) OR 'sodium substitut*':ti,ab,kw OR (('sodium' NEAR/3 'substitute'):ti,ab,kw) OR 'potassium enrich*':ti,ab,kw OR 'potassium rich*':ti,ab,kw OR 'reduce* sodium salt':ti,ab,kw OR (('low' NEAR/3 'sodium' NEAR/3 'salt'):ti,ab,kw) OR (('salt' NEAR/3 'reduction'):ti,ab,kw) OR 'salt replac*':ti,ab,kw OR 'reduc* salt content':ti,ab,kw OR 'reduc* sodium content':ti,ab,kw OR 'reduc* dietary sodium':ti,ab,kw OR 'reduc* dietary salt':ti,ab,kw OR 'reduc* salt intake':ti,ab,kw OR 'reduc* sodium intake':ti,ab,kw OR (('low' NEAR/3 'salt'):ti,ab,kw) OR (('low' NEAR/3 'sodium'):ti,ab,kw) OR (('sodium' NEAR/3 'free'):ti,ab,kw) OR (('discretionary' NEAR/3 'salt'):ti,ab,kw)) AND ('dash diet'/exp OR 'dash diet' OR 'hypertension'/exp OR 'hypertension' OR 'blood pressure'/exp OR 'blood pressure' OR 'cardiovascular disease'/exp OR 'cardiovascular disease' OR 'heart disease'/exp OR 'heart disease' OR 'heart failure'/exp OR 'heart failure' OR 'heart muscle ischemia'/exp OR 'heart muscle ischemia' OR 'cerebrovascular accident'/exp OR 'cerebrovascular accident' OR 'ischemic stroke'/exp OR 'ischemic stroke' OR 'brain hemorrhage'/exp OR 'brain hemorrhage' OR 'subarachnoid hemorrhage'/exp OR 'subarachnoid hemorrhage' OR 'normotens*':ti,ab,kw OR 'hypertens*':ti,ab,kw OR 'blood pressure':ti,ab,kw OR 'cardiovascular disease*':ti,ab,kw OR 'heart disease*':ti,ab,kw OR 'heart failure':ti,ab,kw OR 'myocardial infarct*':ti,ab,kw OR 'heart attack*':ti,ab,kw OR 'ischemic heart':ti,ab,kw OR 'ischaemic heart':ti,ab,kw OR 'stroke':ti,ab,kw OR 'intracerebral hemorrhag*':ti,ab,kw OR 'intracerebral haemorrhag*':ti,ab,kw OR 'cerebral hemorrhag*':ti,ab,kw OR 'cerebral haemorrhag*':ti,ab,kw OR 'intracranial hemorrhag*':ti,ab,kw OR 'intracranial haemorrhag*':ti,ab,kw OR 'cranial hemorrhag*':ti,ab,kw OR 'cranial haemorrhag*':ti,ab,kw OR 'subarachnoid hemorrhag*':ti,ab,kw OR 'subarachnoid haemorrhag*':ti,ab,kw OR 'brain infarct*':ti,ab,kw OR 'cerebral infarct*':ti,ab,kw OR 'cerebrovascular accident*':ti,ab,kw OR 'death'/exp OR 'death' OR 'death':ti,ab,kw) AND ('meta analysis'/de OR 'systematic review'/de OR 'meta analysis topic'/de OR 'systematic review topic'/de) AND [embase]/lim NOT ([embase]/lim AND [medline]/lim) | 02/04/2026 | 228 |
| [Global Health (EBSCO)](https://offcampus.lib.washington.edu/login?url=https://search.ebscohost.com/login.aspx?authtype=ip,uid&profile=ehost&defaultdb=lhh) | (((MH "Potassium, dietary+") OR (MW "potassium dietary") OR (MW "potassium chloride") OR (MH "potassium chloride+") OR (TI "potassium chloride" OR AB "potassium chloride") OR (MW "potassium citrate") OR (MH "potassium citrate+") OR (TI "potassium citrate" OR AB "potassium citrate") OR "potassium salt[tiab:~2]" OR (TI "KCl salt" OR AB "KCl salt") OR (TI "salt substitut*" OR AB "salt substitut*") OR "salt substitute[tiab:~2]" OR "salt substitution[tiab:~2]" OR (TI "sodium substitut*" OR AB "sodium substitut*") OR "sodium substitute[tiab:~2]" OR (TI "potassium enrich*" OR AB "potassium enrich*") OR (TI "potassium rich*" OR AB "potassium rich*") OR (TI "reduce* sodium salt" OR AB "reduce* sodium salt") OR "low sodium salt[tiab:~2]" OR "salt reduction[tiab:~2]" OR (TI "salt replac*" OR AB "salt replac*") OR (TI "reduc* salt content" OR AB "reduc* salt content") OR (TI "reduc* sodium content" OR AB "reduc* sodium content") OR (TI "reduc* dietary sodium" OR AB "reduc* dietary sodium") OR (TI "reduc* dietary salt" OR AB "reduc* dietary salt") OR (TI "reduc* salt intake" OR AB "reduc* salt intake") OR (TI "reduc* sodium intake" OR AB "reduc* sodium intake") OR "low salt[tiab:~2]" OR "low sodium[tiab:~2]" OR "sodium free[tiab:~2]" OR "discretionary salt[tiab:~2]" ) AND ((MH "Dietary Approaches To Stop Hypertension+") OR (MH Hypertension+) OR (MH "Blood Pressure+") OR (MH "Cardiovascular Diseases+") OR (MH "Heart Diseases+") OR (MH "Heart Failure+") OR (MH "Myocardial Ischemia+") OR (MH Stroke+) OR (MH "Ischemic Stroke+") OR (MH "Intracranial Hemorrhages+") OR (MH "Cerebral Hemorrhage+") OR (MH "Subarachnoid Hemorrhage+") OR (TI normotens* OR AB normotens*) OR (TI hypertens* OR AB hypertens*) OR (TI "blood pressure" OR AB "blood pressure") OR (TI "cardiovascular disease*" OR AB "cardiovascular disease*") OR (TI "heart disease*" OR AB "heart disease*") OR (TI "heart failure" OR AB "heart failure") OR (TI "myocardial infarct*" OR AB "myocardial infarct*") OR (TI "heart attack*" OR AB "heart attack*") OR (TI "ischemic heart" OR AB "ischemic heart") OR (TI "ischaemic heart" OR AB "ischaemic heart") OR (TI stroke OR AB stroke) OR (TI "intracerebral hemorrhag*" OR AB "intracerebral hemorrhag*") OR (TI "intracerebral haemorrhag*" OR AB "intracerebral haemorrhag*") OR (TI "cerebral hemorrhag*" OR AB "cerebral hemorrhag*") OR (TI "cerebral haemorrhag*" OR AB "cerebral haemorrhag*") OR (TI "intracranial hemorrhag*" OR AB "intracranial hemorrhag*") OR (TI "intracranial haemorrhag*" OR AB "intracranial haemorrhag*") OR (TI "cranial hemorrhag*" OR AB "cranial hemorrhag*") OR (TI "cranial haemorrhag*" OR AB "cranial haemorrhag*") OR (TI "subarachnoid hemorrhag*" OR AB "subarachnoid hemorrhag*") OR (TI "subarachnoid haemorrhag*" OR AB "subarachnoid haemorrhag*") OR (TI "brain infarct*" OR AB "brain infarct*") OR (TI "cerebral infarct*" OR AB "cerebral infarct*") OR (TI "cerebrovascular accident*" OR AB "cerebrovascular accident*") OR (MH death+) OR (TI death OR AB death))) AND (((PT "meta analysis") OR (PT "systematic review") OR (MH "meta analysis as topic+") OR (MH "systematic reviews as topic+") OR (SB systematic) OR "systematic review[tiab:~2]" OR "systematically reviewed[tiab:~2]" OR "meta analysis" OR "meta analyses" OR metaanaly* OR "cochrane review" OR "critical review" OR "evidence gap map" OR "integrative review" OR "mapping review" OR "mixed studies review" OR "mixed methods review" OR "evidence synthesis" OR "research synthesis" OR "rapid review" OR "realist review" OR "scoping review[tiab:~2]" OR "state-of-the-art review" OR "systematized review" OR "umbrella review" OR "review of reviews" OR (SO "The Cochrane database of systematic reviews" OR ST "The Cochrane database of systematic reviews" OR IB "The Cochrane database of systematic reviews"))) | 02/04/2026 | 44 |
| [Web of Science](https://www.offcampus.lib.washington.edu/login?url=https://www.webofscience.com/wos/woscc/basic-search) | ((TS=(("potassium dietary" OR "potassium chloride" OR "potassium citrate" OR "potassium salt" OR "KCl salt" OR "salt substitut*" OR "salt substitute" OR "salt substitution" OR "sodium substitut*" OR "sodium substitute" OR "potassium enrich*" OR "potassium rich*" OR "reduce* sodium salt" OR "low sodium salt" OR "salt reduction" OR "salt replac*" OR "reduc* salt content" OR "reduc* sodium content" OR "reduc* dietary sodium" OR "reduc* dietary salt" OR "reduc* salt intake" OR "reduc* sodium intake" OR "low salt" OR "low sodium" OR "sodium free" OR "discretionary salt") )) AND TS=(("Dietary Approaches To Stop Hypertension" OR "Hypertension" OR "Blood Pressure" OR "Cardiovascular Disease" OR "Heart Disease" OR "Heart Failure" OR "Myocardial Ischemia" OR "Stroke" OR "Ischemic Stroke" OR "Intracranial Hemorrhage" OR "Cerebral Hemorrhage" OR "Subarachnoid Hemorrhage" OR normotens* OR hypertens* OR "myocardial infarct*" OR "heart attack*" OR "ischemic heart" OR "ischaemic heart" OR "intracerebral hemorrhag*" OR "intracerebral haemorrhag*" OR "cerebral hemorrhag*" OR "cerebral haemorrhag*" OR "intracranial hemorrhag*" OR "intracranial haemorrhag*" OR "cranial hemorrhag*" OR "cranial haemorrhag*" OR "subarachnoid hemorrhag*" OR "subarachnoid haemorrhag*" OR "brain infarct*" OR "cerebral infarct*" OR "cerebrovascular accident*" OR "death"))) AND TS=(("meta analysis" OR "systematic review" OR "systematically reviewed" OR "meta analysis" OR "meta analyses" OR metaanaly* OR "cochrane review" OR "critical review" OR "evidence gap map" OR "integrative review" OR "mapping review" OR "mixed studies review" OR "mixed methods review" OR "evidence synthesis" OR "research synthesis" OR "rapid review" OR "realist review" OR "scoping review" OR "state-of-the-art review" OR "systematized review" OR "umbrella review" OR "review of reviews")) | 02/12/2026 | 572; 209 after refined to Document Type: Review Articles |
| [Cochrane](http://offcampus.lib.washington.edu/login?url=https://www.cochranelibrary.com/) Systematic Reviews | ("potassium dietary" OR "potassium chloride" OR "potassium citrate" OR "potassium salt" OR "KCl salt" OR "salt substitut*" OR "salt substitute" OR "salt substitution" OR "sodium substitut*" OR "sodium substitute" OR "potassium enrich*" OR "potassium rich*" OR "reduce* sodium salt" OR "low sodium salt" OR "salt reduction" OR "salt replac*" OR "reduc* salt content" OR "reduc* sodium content" OR "reduc* dietary sodium" OR "reduc* dietary salt" OR "reduc* salt intake" OR "reduc* sodium intake" OR "low salt" OR "low sodium" OR "sodium free" OR "discretionary salt") in Title Abstract Keyword AND ("Dietary Approaches To Stop Hypertension" OR "Hypertension" OR "Blood Pressure" OR "Cardiovascular Disease" OR "Heart Disease" OR "Heart Failure" OR "Myocardial Ischemia" OR "Stroke" OR "Ischemic Stroke" OR "Intracranial Hemorrhage" OR "Cerebral Hemorrhage" OR "Subarachnoid Hemorrhage" OR normotens* OR hypertens* OR "myocardial infarct*" OR "heart attack*" OR "ischemic heart" OR "ischaemic heart" OR "intracerebral hemorrhag*" OR "intracerebral haemorrhag*" OR "cerebral hemorrhag*" OR "cerebral haemorrhag*" OR "intracranial hemorrhag*" OR "intracranial haemorrhag*" OR "cranial hemorrhag*" OR "cranial haemorrhag*" OR "subarachnoid hemorrhag*" OR "subarachnoid haemorrhag*" OR "brain infarct*" OR "cerebral infarct*" OR "cerebrovascular accident*" OR "death") in Title Abstract Keyword - in Cochrane Reviews (Word variations have been searched) | 2/12/2026 | 20 |

**Supplementary Table S3. Characteristics of included reviews evaluating potassium-containing low-sodium salt substitutes for primary and secondary prevention of cardiovascular disease.**

| **Review** **(author, year)** | **Review** **type** | **Last search** **date** | **Included studies** **(n; design)** | **Prevention** **status†** | **Population /** **special subgroups** | **Geographic** **scope** | **LSSS formulation /** **comparator** | **Outcomes covered‡** | **Keynote** |
| --- | --- | --- | --- | --- | --- | --- | --- | --- | --- |
| Hernandez 2019 | SR+MA | 31 May 2018 | 21 RCTs; n=7,403 | Primary | Adults; hypertensive, normotensive, prehypertensive, or mixed | 8 countries; China-dominant | KCl-based LSSS (25–49% KCl), some Mg/Ca; regular salt comparator | T1: ACM, detected HTN T2: SBP, DBP, 24-h urinary Na/K/Ca | Pre-SSaSS; primary-prevention only; some non-KCl studies |
| Jafarnejad 2020 | SR+MA | Dec 2018 | 10 studies/11 comparisons; n=1,119 | Primary | Adults with stage 2 HTN | China plus Europe/Brazil | NaCl/KCl blends (~50:50 to 65:30), some Mg/Ca; regular salt comparator | T2: SBP, DBP | HTN-only; pre-SSaSS; 1 chitosan trial in source pool |
| Jin 2020 § | SR+MA | Oct 2019 | 24 RCTs overall; SS subgroup 13 studies, n=5,653 | Mixed/unclear | Chinese populations; adults, children, and high-risk groups | China only | K-based SS subgroup; regular salt comparator | T2: SBP, DBP | Broader salt-reduction review; SS subgroup extracted |
| Yin 2022 | SR+MA | 31 Aug 2021 | 21 RCTs; n=31,949 | Mixed/unclear | Adults; mixed BP status; some prior stroke/TIA | Multi-region; China-dominant | KCl-based SS (NaCl 33–75%; KCl 25–65%), some Mg; regular salt comparator | T1: ACM, CVD mort, MACE T2: SBP, DBP, 24-h urinary Na/K T3: serum K, hyperkalaemia | First review to pool clinical outcomes; SSaSS-dominant |
| Brand 2022 | Cochrane SR+MA | 18 Aug 2021 | 26 RCTs; n=35,053 | Mixed/unclear | Adults predominantly; mixed BP/CVD risk; 1 child trial | Global/multi-region; majority Asian settings | Potassium-containing LSSS with variable K content; regular salt or no intervention | T1: stroke, ACM, CVD mort, MACE T2: SBP, DBP, 24-h urinary Na/K T3: serum K, hyperkalaemia, creatinine, SAE | Most comprehensive pairwise review; some non-KCl/unclear arms |
| Tsai 2022 | SR+MA | Mar 2022 | 23 RCTs; n=32,073 | Mixed/unclear | Adults; 13 HTN-only trials plus mixed BP populations | 14 countries; China-dominant | Heterogeneous K-based SS; regular salt comparator | T1: ACM, CVD mort T2: SBP, DBP, urinary Na/K, Na:K T3: serum K | Only review with pooled urinary Na:K ratio |
| Aliasgharzadeh 2022 § | SR+MA | Apr 2022 | 40 articles/50 arms (35/44 in MA); n=33,320 | Mixed/unclear | Children and adults; mixed BP/CVD risk | Multi-region/global | Salt-substitute sub-analysis within broader salt-reduction review; mostly regular salt/usual care | T2: SBP, DBP | SS pooled estimate not clean LSSS-only (chitosan contamination; SSaSS-dominant) |
| Greenwood 2024 | SR+MA | 23 Aug 2023 | 16 RCTs; n=35,251 | Mixed/unclear | Adults ≥18 years; long-term trials (≥6 mo); mixed BP/CVD risk | China/Taiwan plus Peru and Europe | K-based SS; explicit formulations 25–66% KCl, some Mg/Ca; regular salt/usual care | T1: ACM, CVD mort, MACE T2: SBP, DBP, 24-h urinary Na/K T3: serum K, SAE | Long-term review; key anchor for hard outcomes |
| Prommas 2026 § | SR+NMA | 28 Feb 2025 | 42 studies/46 trials; n=46,771 overall | Mixed/unclear | Adults with elevated BP or hypertension | Multi-region; China-dominant | Behavioural NMA with a salt-substitute node; no intervention or health education comparators | T2: SBP only | SS estimate contaminated by chitosan trials and dominated by SSaSS |
| Nguyen 2026 § | SR+NMA | Mar 2025 | 50 RCTs overall; SS node 1 trial, n=20,995 | Mixed/unclear | High-risk stroke-prevention populations overall; SS node from SSaSS | Global NMA; SS node from China | Salt-substitute node vs regular salt; formulation not detailed in source review | T1: total, fatal, haemorrhagic, fatal haemorrhagic stroke | Broader stroke-prevention NMA; all SS estimates from a single trial |
| Lai 2026 | SR+NMA | NR | 34 RCTs; n=37,063 | Mixed/unclear | Adults ≥18 years; formulation-specific network; mixed-risk populations | 15 countries; China-dominant | Published formulation nodes (K-salt, moderate-K, high-K, low-Na); regular salt, other substitutes, or no intervention | T1: ACM, CVD mort, non-fatal CVD T2: SBP, DBP, 24-h urinary Na/K T3: serum K, AE withdrawals | First formulation-specific NMA |

Reviews are ordered by last search date (oldest first). Prevention status was classified by established cardiovascular disease at baseline rather than by hypertension status alone. Broader salt-reduction reviews that reported an extractable potassium-containing low-sodium salt substitute subgroup or network node are indicated in the notes. Outcome coverage is summarised at review level; extractable pooled estimates are detailed in Supplementary Table S3.

**Abbreviations:** ACM, all-cause mortality; AE, adverse event; CVD, cardiovascular disease; CVD mort, cardiovascular mortality; DBP, diastolic blood pressure; HTN, hypertension; LSSS, low-sodium salt substitute; MA, meta-analysis; MACE, major adverse cardiovascular events; NMA, network meta-analysis; NR, not reported; RCT, randomised controlled trial; SAE, serious adverse event; SBP, systolic blood pressure; SS, salt substitute.

† Prevention status was classified by established cardiovascular disease at baseline, not by hypertension status alone.

‡ Outcome coverage is summarised at review level; Supplementary Table S3 indicates whether pooled estimates were extractable on a comparable scale.

§ Reviews not specific to potassium-containing LSSS contributed an extractable salt-substitute subgroup or network node only.

**Supplementary Table S4. Outcome-by-review evidence map across the prespecified three-tier framework.**

| **Outcome** | **Hernandez**  **2019** | **Jafarnejad**  **2020** | **Jin**  **2020** | **Yin**  **2022** | **Brand**  **2022** | **Tsai**  **2022** | **Aliasgharzadeh**  **2022** | **Greenwood**  **2024** | **Prommas**  **2026** | **Nguyen**  **2026** | **Lai**  **2026** |
| --- | --- | --- | --- | --- | --- | --- | --- | --- | --- | --- | --- |
| **Tier 1: Clinical outcomes** | | | | | | | | | | | |
| Major CVD events (composite) | — | — | — | **Y** | — | — | — | **Y** | — | — | — |
| Non-fatal CVD events | — | — | — | — | — | — | — | — | — | — | **A** |
| Stroke (non-fatal) | — | — | — | — | **Y** | — | — | — | — | — | — |
| Fatal stroke | — | — | — | — | — | — | — | — | — | **Y** | — |
| Hemorrhagic stroke | — | — | — | — | — | — | — | — | — | **Y** | — |
| Fatal hemorrhagic stroke | — | — | — | — | — | — | — | — | — | **Y** | — |
| Detected hypertension | **N** | — | — | — | — | — | — | — | — | — | — |
| CVD mortality | — | — | — | **Y** | **Y** | **Y** | — | **Y** | — | — | **A** |
| All-cause mortality | **N** | — | — | **Y** | **Y** | **Y** | — | **Y** | — | — | **A** |
| **Tier 2: Intermediate/biomarker outcomes** | | | | | | | | | | | |
| Systolic blood pressure (SBP) | **Y** | **Y** | **Y** | **Y** | **Y** | **Y** | **Y** | **Y** | **Y** | — | **Y** |
| Diastolic blood pressure (DBP) | **Y** | **Y** | **Y** | **Y** | **Y** | **Y** | **Y** | **Y** | — | — | **Y** |
| 24-h urinary sodium excretion | **Y** | — | — | **Y** | **N** | **Y** | — | **Y** | — | — | **Y** |
| 24-h urinary potassium excretion | **Y** | — | — | **Y** | **Y** | **Y** | — | **Y** | — | — | **Y** |
| Urinary Na:K ratio | — | — | — | — | — | **Y** | — | — | — | — | — |
| **Tier 3: Safety outcomes** | | | | | | | | | | | |
| Serum potassium | — | — | — | **Y** | **Y** | **Y** | — | **Y** | — | — | **Y** |
| Hyperkalaemia (events) | — | — | — | — | **Y** | — | — | — | — | — | — |
| Serious adverse events (SAE) | — | — | — | — | — | — | — | **Y** | — | — | — |
| Serum creatinine | — | — | — | — | **N** | — | — | — | — | — | — |
| AE-related withdrawals | — | — | — | — | — | — | — | — | — | — | **Y** |

Matrix showing which included reviews reported extractable pooled estimates for Tier 1 clinical outcomes, Tier 2 intermediate/biomarker outcomes and Tier 3 safety outcomes. Y = extractable pooled estimate; N = outcome reported but not pooled or pooled estimate not extractable on the target scale; A = absolute effects only; — = not reported. Nguyen 2026 salt-substitute estimates are from a single direct trial (SSaSS). Lai 2026 reports Tier 1 clinical outcomes as absolute effects rather than directly comparable RR/HR. Aliasgharzadeh 2022 and Prommas 2026 include published salt-substitute estimates that are not fully protocol-concordant because ineligible chitosan-containing trials were included in the source review estimates. Brand 2022 did not pool 24-h urinary sodium because heterogeneity was high (I² = 91%); serum creatinine was reported but non-significant. Hernandez 2019 reported non-significant pooled estimates for all-cause mortality and detected hypertension.

**Supplementary Table S5. Primary-study overlap matrix used to calculate corrected covered area across included reviews.**

| Number of columns (number of reviews) | c | 11 |
| --- | --- | --- |
| Number of rows (number of index publications) | r | 40 |
| Number of included primary studies (including double counting) | N | 154 |
| Covered area | N/(rc) | 35.00% |
| Corrected covered area | (N-r)/(rc-r) | 28.50% |
| Interpretation of overlap | **Very High overlap** | |
| Corrected covered area   (adjusting by structural zeros) | (N-r)/(rc-r-X) | 28.50% |
| N° of non-overlapped primary studies | In 1 SR | 10 |
| Number of overlapped primary studies | In 2 SRs | 4 |
|  | In 3 SRs | 6 |
|  | In 4 SRs | 4 |
|  | In 5 SRs | 5 |
|  | In 6 SRs | 5 |
|  | In 7 SRs | 3 |
|  | In 8 SRs | 1 |
|  | In 9 SRs | 2 |
|  | In 10 SRs | 0 |
|  | In 11 SRs | 0 |

A citation matrix of protocol-eligible primary studies were summarised across included reviews with rows representing unique primary studies and columns representing included reviews; filled cells indicate that a primary study was included in the corresponding review. A matrix was used to calculate the overall corrected covered area as a measure of primary-study overlap. Studies reported within otherwise eligible reviews but not meeting the umbrella review intervention definition were identified separately and excluded from the primary CCA calculation. Abbreviation: CCA, corrected covered area. N°, Number.

**Supplementary Table S6. Formulation heterogeneity of potassium-containing low-sodium salt substitutes across included reviews**

| **Review** | **Published KCl content** | **Reported formulations / categories** | **Formulation analysis** | **Main formulation-related finding** |
| --- | --- | --- | --- | --- |
| **Hernandez 2019** | 25–49% KCl | 14 reported trial formulations | Meta-regression (NaCl content vs SBP) | No significant association between NaCl content and blood-pressure effect was reported. |
| **Jafarnejad 2020** | 25–50% KCl | 8 explicit formulations | None | No formal formulation-stratified analysis; published BP estimate includes one non-KCl chitosan study. |
| **Jin 2020** | 25–50% KCl | 13 salt-substitute study populations (explicit ratios reported for 8) | Meta-regression (salt-reduction strategy type) | Salt-substitute type was not significantly associated with SBP or DBP (p=0.47 and p=0.06). |
| **Yin 2022** | 25–65% KCl | 21 trials | Meta-regression (NaCl and KCl proportion, continuous) | Each 10% lower NaCl content was associated with lower SBP (MD −1.53 mmHg, 95% CI −3.02 to −0.03; p=0.045). |
| **Brand 2022** | 25–66% KCl | 10 trials ≥30% KCl; 12 <30%; 4 unclear | Prespecified subgroup and univariable meta-regression | No important subgroup differences in SBP or DBP by KCl proportion were reported. |
| **Tsai 2022** | 25–65% KCl | 23 trials | None | No formal formulation-stratified analysis. |
| **Aliasgharzadeh 2022** | 25–30% KCl explicitly reported; some ratios NR | 11 articles / 14 salt-substitute trial arms | None | Published salt-substitute subgroup includes non-KCl chitosan arms and excludes some eligible potassium-containing trials; formulation effects were not analysed separately. |
| **Greenwood 2024** | 25–66% KCl | 16 trials; KCl subgroup analysis available for 11 | Prespecified subgroup (≥30% vs <30% KCl) | SBP reduction was larger with ≥30% KCl (MD −7.88, 95% CI −9.45 to −6.30; k=4) than with <30% KCl (MD −3.51, 95% CI −4.91 to −2.10; k=7); p-interaction <0.01. |
| **Prommas 2026** | NR at review level | Salt-substitute treated as one NMA node (14–15 trials across salt-substitute comparisons) | None | Trial formulations were not reported at node level; published SBP node includes non-KCl chitosan trials. |
| **Nguyen 2026** | 25% KCl / 75% NaCl | 1 direct trial (SSaSS) | None | All potassium-containing LSSS evidence reflects one formulation only. |
| **Lai 2026** | 3–66% KCl across published classifications | 34 trials; overlapping NMA categories | NMA by published formulation categories | Published SBP estimates were greatest for high-potassium salt substitutes (50–65% KCl: MD −7.54, 95% CI −11.63 to −3.44), followed by moderate-potassium salt substitutes (25–40% KCl: MD −4.64, 95% CI −6.05 to −3.24), low-sodium salt substitutes (60–79% NaCl: MD −4.39, 95% CI −6.02 to −2.75), and the broader K-salt category (MD −4.00, 95% CI −5.67 to −2.32); published categories overlap and are not mutually exclusive. |

Published formulation categories are reported as stated by the source review and do not necessarily represent mutually exclusive protocol-specific NaCl–KCl-only nodes. Several reviews also included potassium-containing formulations with magnesium, calcium, citrate, lysine, folate, flavorings, or other co-constituents. Lai 2026 reported overlapping published network categories (K-salt, moderate-potassium, high-potassium, and low-sodium salt substitutes). A single trial could contribute to more than one published category. Some published pooled estimates were partially extractable because the source review included non-KCl chitosan trials (Allaert 2013 and/or Allaert 2017) that could not be separated from the reported summary estimate. **Abbreviations:** BP, blood pressure; DBP, diastolic blood pressure; KCl, potassium chloride; LSSS, low-sodium salt substitute; NMA, network meta-analysis; NR, not reported; SBP, systolic blood pressure; SSaSS, Salt Substitute and Stroke Study.
